# Editing stem cell genomes at scale to measure variant effects in diverse cell and genetic contexts

**DOI:** 10.1101/2025.11.12.25340127

**Authors:** Shawn Fayer, Riddhiman K. Garge, Melissa Hopkins, Clayton E. Friedman, Abby V. McGee, Joshua Rico, Rachel L. Powell, Evan McDermot, Nahum T. Smith, Sriram Pendyala, Marcy E. Richardson, Erica D. Smith, B. Monica Bowen, Rebecca Resnick, Pankhuri Gupta, Andrew B. Stergachis, Casey Gifford, Sudarshan Pinglay, Kai-Chun Yang, Douglas M. Fowler, Lea M. Starita

**Author notes:** Corresponding authors, L. Starita, D. Fowler and S. Fayer.

## Abstract

Multiplexed assays of variant effect (MAVEs) systematically measure variant function but have been limited to cancer cell lines rather than disease-relevant cell types. We developed saturation genome editing in human iPSCs (iPSC-SGE) to introduce variant libraries into a single allele of a target gene while programming the genetic background of the second allele, enabling variant assessment across differentiated cell types and genetic contexts at scale. We edited 1,137 variants into *MYBPC3* and measured protein abundance in cardiomyocytes and cardiac organoids, accurately identifying pathogenic variants, and resolving variants of uncertain significance. Highlighting the importance of genetic context, we edited 437 *POLG* variants in two genetic backgrounds and identified loss-of-function and dominant-negative variants. Finally, we illuminate a path for scaling iPSC-SGE by identifying 443 disease genes essential for iPSC or iPSC-derived neuron growth. iPSC-SGE enables systematic assessment of variants in specialized human cell types, advancing MAVEs to empower genomic medicine.

## Introduction

The effects of genetic variation are closely tied to their cell and genetic context. Variants often cause disease by impacting a particular cell type at a particular point in development. For example, many genes associated with cardiomyopathies encode structural components of the sarcomere and are exclusively expressed in cardiomyocytes. Likewise, variant effects often depend on genetic context, most fundamentally with respect to the interaction between two copies of the same gene. For a given condition, the specific combination of alleles often defines phenotypic severity. Thus, accurate measurement of variant functional effects is dependent on our ability to express variants in differentiated cell types in the context of different genetic backgrounds.

Editing variants directly into the genomes of cells has enabled the assessment of variant effects on splicing, cell growth, and other phenotypes at scale. These multiplexed assays of variant effect (MAVEs) with endogenous genome editing have been instrumental in understanding variant impacts on tumor suppressor function [1–9] and tackling variants of uncertain significance (VUS), which arise when a genetic test reveals a variant but there is not enough information to interpret the variant as pathogenic or benign. VUS cannot be used to diagnose disease or guide therapy and there are over 800,000 missense VUS in well established disease genes currently in ClinVar (**Figure 1A**)[10,11]. Unfortunately, most current scaled endogenous genome editing strategies employ cancer-derived or non-human cell lines and thus cannot readily be applied to most clinically relevant genes because these genes encode proteins which function in differentiated cell types (**Figure 1B**). Moreover, editing-based MAVEs have largely been deployed in haploid cells which cannot consider the combined effect of both alleles for recessive disorders, nor measure dominant negative phenotypes.

**Figure 1:**
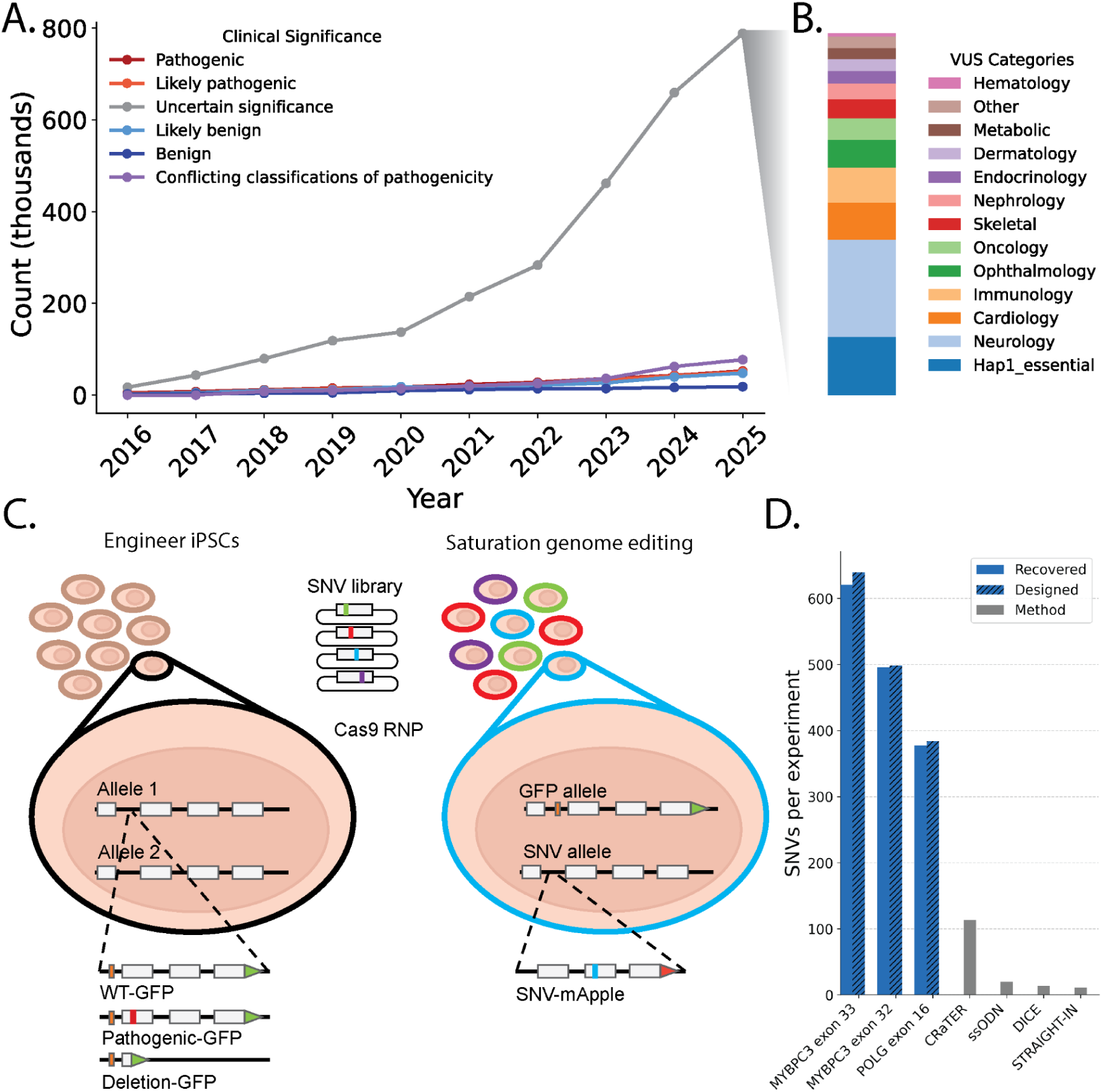
Saturation genome editing (SGE) in iPSCs. **A**) Unique ClinVar missense variants plotted annually by clinical significance. Variants are included from genes in the GenCC database[11] with at least moderate evidence of gene-disease association. Pathogenic/Likely pathogenic and Benign/Likely benign variants were included in the “Likely” category only. **B**) A stacked barplot of VUS colored by genetic testing panel indication. **C**) Schematic of the iPSC-SGE editing workflow; the background allele is first edited to fuse a GFP to the target gene and insert a small deletion (represented by the orange box in the cell on the right) to prevent cutting when the SNV library is introduced. SNVs are then edited into the second allele fused to mApple and an antibiotic selection cassette. **D**) The actual and expected number of SNVs introduced into exon 16 of *POLG* and exons 32 and 33 of *MYBPC3* with iPSC-SGE compared to the maximum number of variants from other iPSC multiplexed editing studies.

Human induced pluripotent stem cells (iPSCs) offer a solution to these limitations by enabling variant assessment in diploid contexts across differentiated cell types. iPSCs preserve the genetic architecture necessary for measuring recessive and dominant negative phenotypes while providing access to cell types where disease-relevant genes are expressed and function. Moreover, because regulatory sequences are left intact, endogenous editing in iPSCs maintains physiologic expression levels during differentiation and avoids silencing often observed with transgenic approaches in differentiated cells. However, large-scale variant effect experiments in iPSCs have been limited by inefficient[12–14] or non-specific editing approaches[15].

Here, we develop saturation genome editing in human iPSCs (iPSC-SGE), a method that leverages homology-directed repair (HDR) to replace endogenous gene segments while simultaneously installing selectable markers and reporters. iPSC-SGE enables assessment of variant effects at the endogenous locus across cell types and genetic contexts. We show that iPSC-SGE supports scaled experiments by editing a total of 1,574 variants into two genes, a ∼14-fold increase compared to the state of the art [12]. We edited 1,137 variants into two exons of *MYBPC3*, where pathogenic variants are the most common cause of familial hypertrophic cardiomyopathy. We differentiated *MYBPC3-*edited iPSCs into cardiomyocytes and measured MYBPC3 variant protein abundance, accurately identifying pathogenic variants and enabling resolution of 67% of VUS. We also created *MYBPC3* variant-bearing cardiac organoids, recovered and scored all variants, demonstrating the feasibility of MAVEs in 3D organoids. We mapped essential binding interface residues required for MYBPC3 integration into the cardiac sarcomere and show that variant effect predictors perform poorly at these positions further highlighting the value of measuring variant effects in cell type context. We edited 437 variants into *POLG*, which encodes the DNA polymerase required for replicating the mitochondrial genome. Pathogenic *POLG* variants are a leading cause of inherited mitochondrial depletion syndromes, and are inherited in both autosomal dominant and recessive manners. Leveraging iPSC-SGE to measure *POLG* variant effects on cell growth in iPSCs with multiple *POLG* background alleles, we identify 117 loss of function (LOF) and 23 dominant negative variants. Finally, we illustrate the generalizability of our approach by identifying 443 disease-relevant target genes for iPSC-SGE with a growth phenotype in iPSC or neurons, demonstrating efficient, one-step, biallelic editing, and developing a strategy to replace whole genes spanning tens of kilobases. iPSC-SGE enables multiplexed assays for genes best phenotyped in specialized cell types and facilitates variant effect mapping in multiple, endogenous genetic backgrounds.

## Results

### Saturation genome editing of over 1,500 variants into induced pluripotent stem cells (iPSC) with iPSC-SGE

To harness the power of iPSCs to assess variant effects in differentiated cell types and in the diploid genetic context we developed iPSC-SGE, a two-step strategy to edit a library of variants into a target gene on a specific genetic background. In the first step, one allele of the target gene, referred to as the background allele hereafter, is modified to install an antibiotic resistance cassette to enable selection for integrants, a terminal GFP fusion to select for in-frame edits and a specific genetic background (e.g. reference, null, or a pathogenic missense variant) (**Figure 1C, Supplementary Figure 1A**). iPSC-SGE repair templates are designed with a 100-200 base intronic deletion directly adjacent to the Cas9 cut site to enrich HDR events that contain the full repair template and prevent cleavage of the background allele in subsequent steps. Antibiotic resistant, GFP positive cells harboring in-frame background allele edits are isolated, with the addition of CRISPR activation to induce transient expression of the target gene if it is not natively expressed in iPSCs [12]. We isolated multiple clonal lines for each target gene, sequence verified each background allele with nanopore sequencing, and validated differentiation potential. In the second step, HDR is used to integrate the variant library specifically into the second allele, targeting the region that was deleted in the background allele. Variant library repair templates have mApple directly fused or co-expressed with the target gene to enable selection of on-target edits.

We applied iPSC-SGE to *MYBPC3* and *POLG*, introducing exon scale saturation variant libraries totaling 1,574 single nucleotide variants (SNVs). We edited *MYBPC3* saturation libraries into WTC-11 cells bearing a wild type background allele because pathogenic variants have an autosomal dominant inheritance pattern (**Figure 1D**). Our libraries spanned exons 32 and 33 encoding the *MYBPC3* C10 domain, a hotspot for pathogenic missense variants [16]. We also varied flanking intronic sequences (**Supplementary Figure 1B**). Edited iPSCs contained 1,137 (100%) of the designed SNVs. We edited a *POLG* saturation library spanning exon 16 and flanking intronic sequence into iPSCs with both null and partial function (W748S) background alleles to assess both loss of function (LOF) and dominant negative phenotypes (**Supplementary Figure 2A, B**). *POLG* edited iPSCs contained 437 (99.8%) of the 438 designed SNVs. This represents a ∼14-fold increase in the scale of variant libraries edited into human stem cells in a single experiment (**Figure 1D**) [13,17–19] libraries on multiple backgrounds, a feat not previously achieved.

### iPSC-SGE identifies LOF variants in *MYBPC3* in cardiomyocytes and cardioids

*MYBPC3* encodes cardiac myosin binding protein C3, which is exclusively expressed in cardiomyocytes and serves as a key structural component of the sarcomere, modulating cardiomyocyte contraction. Autosomal dominant pathogenic variants in *MYBPC3* are the leading cause of familial hypertrophic cardiomyopathy (HCM) [20,21]. The *MYBPC3* iPSC libraries we created enable phenotyping of *MYBPC3* variants after differentiation to cardiomyocytes. An established pathogenic mechanism for missense variants in the C10 domain is reduced incorporation into the sarcomere due to loss of protein stability and, consequently, cellular abundance [22]. To model dominant cardiomyopathy associated variants, we differentiated edited iPSCs to cardiomyocytes following a modified monolayer, small molecule directed differentiation protocol [12] (**Figure 2A**). The MYBPC3-GFP fusion encoded by the reference background allele had the anticipated sarcomere localization pattern in all cardiomyocytes. The variant MYBPC3-mApple fusions, encoded by the SNV alleles, had phenotypes ranging from wild type abundance and normal sarcomere localization to loss of abundance with a dim and diffuse pattern (**Supplementary Figure 3**).

**Figure 2:**
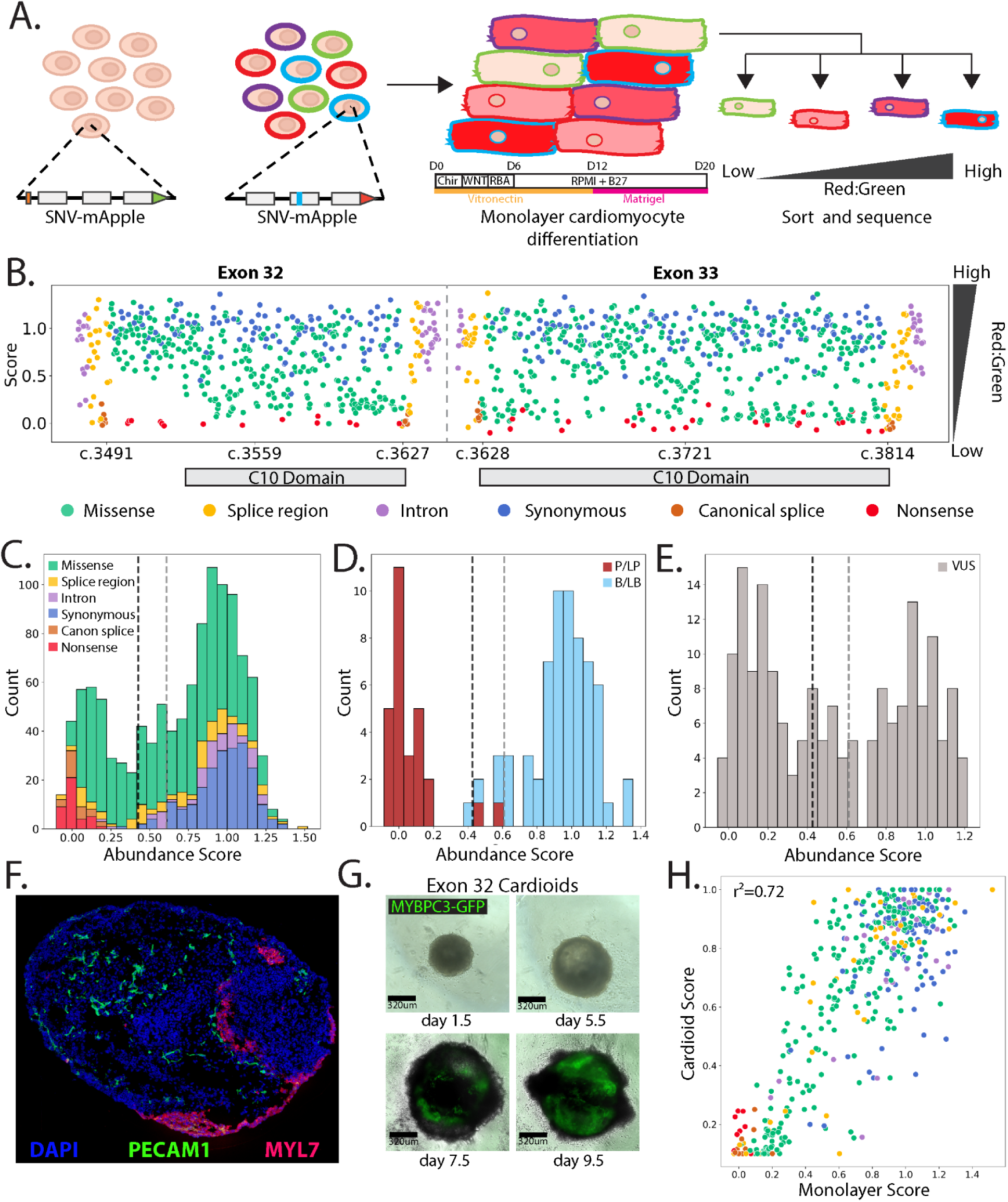
Measuring the effect of *MYBPC3* variants on protein abundance in cardiomyocytes. **A**) Schematic of the *MYBPC3* genomic locus shows a GFP-fused to the reference allele and the mApple fused to the SNV library edited into the second allele. Edited iPSCs are differentiated into cardiomyocytes using a monolayer protocol [12] and cells are sorted according to the mApple/GFP ratio. **B**) Abundance scores for MYBPC3 variants by transcript position with SNVs colored by molecular consequence. The bases encoding the C10 domain are labeled. **C**) Histogram of MYBPC3 abundance scores colored by molecular consequence. Vertical dotted lines represent the thresholds for the functional classes. **D**) Histogram of MYBPC3 abundance scores for ClinVar variants colored by classification. **E**) Histogram of MYBPC3 abundance scores for ClinVar VUS. **F**) Cardioid at day 10.5 of differentiation, sectioned and stained for nuclei (DAPI; blue), PECAM1 (green) and MYL7 (magenta). **G**) Cardioids expressing an MYBPC3-GFP fusion. **H**) Abundance scores for MYBPC3 variants from the 2D monolayer cardiomyocytes vs. 3D cardioids colored by molecular consequence.

To quantify the abundance of each MYBPC3-mApple variant relative to wild type MYBCP3-GFP, we sorted cardiomyocytes into one of four bins based on the mApple to GFP ratio. Variant abundance scores were calculated by taking the weighted average of the variant’s frequency in each bin and normalized across exons by setting the median of the synonymous distributions to 1 and the median of the nonsense distributions to 0 [23] (**Figure 2A).** We classified variants as normal, low, or indeterminate abundance using the distribution of synonymous variants.

Abundance scores for the 1,137 SNVs were bimodally distributed, with nonsense and canonical splice site variants separating from synonymous and intronic variants (**Figure 2B,C**). All nonsense and canonical splice site variants were low abundance along with 219 missense and 16 intronic splice region variants (**Figure 2B and 2C**). Our abundance scores were concordant with variant classifications in the ClinVar database, with 26 of the 28 pathogenic or likely pathogenic (PLP) variants scoring as low abundance and two as indeterminate. 59 of the 63 benign or likely benign (BLB) variants were normal abundance, three were indeterminate, and one was low abundance (**Figure 2D**). 77 of the 178 VUS in this region were low abundance, 22 were indeterminate, and 79 were normal abundance (**Figure 2E, Supplementary Table 1**).

*In vivo*, cardiomyocytes are embedded in the complex tissue of the heart among additional cell types, and subject to significant forces that relate to the pathogenesis of cardiomyopathy. To recapitulate these interactions, cardiac organoids (cardioids) have been employed for drug screening[24,25] and to assess the effects of single variants[26,27]; however, library-scale variant effect experiments have not been performed. Leveraging our ability to create precisely defined variant libraries, we generated cardioids from iPSCs harboring the MYBPC3 exon 32 variant library. These cardioids consist of cardiomyocytes (MYL7 **(Figure 2F**), MYBPC3-GFP (**Figure 2G**)), endothelial cells (PECAM1 (**Figure 2F**)), and other biologically relevant cardiac cell types [28]. We dissociated the cardioids and sorted single cells into GFP positive/mApple negative, and GFP positive/mApple positive bins to generate MYBPC3 cardioid abundance scores. We detected all 498 exon 32 SNVs in cardioids and the cardioid-derived abundance scores were highly correlated with monolayer-derived scores (**Figure 2H**). Together, these results demonstrate that MYBPC3 abundance does not appear to depend strongly on tissue context and that unlike other organoid models, cardioids do not pass through a severe bottleneck limiting the number of cells harboring variants, making them amenable to MAVEs[29].

### iPSC-SGE reveals context-dependent structural features of MYBPC3

Despite decades of research studying *MYBPC3*’s roles in hypertrophic cardiomyopathy, much remains unknown about how pathogenic *MYBPC3* variants cause disease. As a result, clinicians are hesitant to classify missense variants as pathogenic, perpetuating the VUS problem for this gene. MYBPC3 binds actin and myosin during contraction and acts as a brake, limiting contractile force[30–33]. The C10 domain is a C-terminal Ig-like domain that binds to the MYH7 light meromyosin (LMM) tails, anchoring MYBPC3 to the myosin filament [34] [35]. Five charged residues at positions R1205, D1220, D1224, R1241, and K1242 have been shown to be essential for MYH7 binding in a biochemical binding assay[36]. Recent structural analysis has shown that C10 binding is modulated by electrostatic interactions of the negatively charged MYH7 LMM tail residues and the positively charged C10 residues[37].

Our functional data reinforces the importance of this electrostatic interaction at the interface between the C10 domain and the MYH7 LMM tails. The majority of low abundance missense variants in the C10 domain occur at buried hydrophobic residues and likely disrupt Ig-like domain folding (**Figure 3A, B**). However, low abundance missense variants that occur on the surface are enriched at the MYH7 LMM interface (**Figure 3A, B; Supplementary Figure 4A**). Each of the nine positively charged residues with close physical contact to the MYH7 LMM had variants that reduced MYBPC3 abundance (**Supplementary Figure 4A, B**) consistent with the essential role of R1205, R1241, and K1242 for myosin binding.

**Figure 3:**
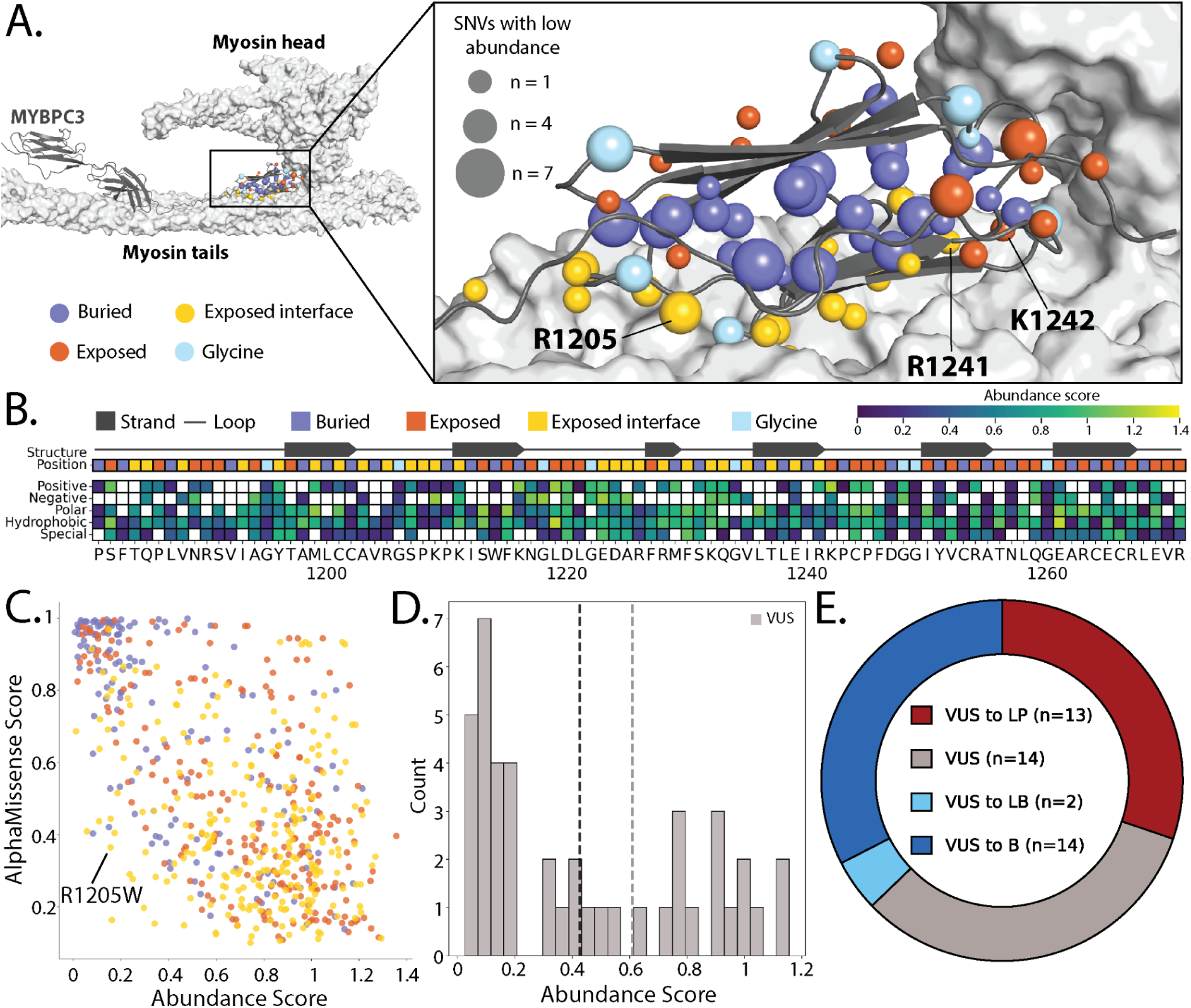
MYBPC3 abundance scores are concordant with structural features and have high clinical utility. **A**) The structure of MYBPC3 on a myosin filament (left) and zoomed-in (right) (pdb_00008g4l)[37]. The MYBPC3 backbone is grey with the C10 domain colored by indicated features. The number of low abundance SNVs at each amino acid is depicted by the size of the sphere at the beta-carbon. **B**) A heatmap of mean abundance scores for amino acid substitutions grouped by chemical properties for each codon position. Beta strands are mapped above and each amino acid position is colored by indicated structural features. **C**) A scatter plot of abundance scores vs. AlphaMissense scores[61] colored by molecular consequence. **D**) A histogram of VUS from Ambry Genetics and the University of Washington Medical Center[62]. **E**) A donut plot of VUS that could be reclassified with addition of functional evidence.

In addition to mapping MYBPC3 residues critical for sarcomere integration, our functional scores offer insights into the limitations of variant effect predictors for structural proteins. While *MYBPC3* iPSC-SGE and predictors are overall in agreement (Pearson’s R 0.42-0.67 and **Supplementary Figure 5**), there are important differences. Variant effect predictors for *MYBPC3* predict more variants as deleterious compared to our functional data. iPSC-SGE-derived abundance score and AlphaMissense prediction discordance is greatest at exposed residues (**Figure 3C**) with AlphaMissese predicting many normal abundance variants to be deleterious at residues facing away from the MYH7 interface (**Figure 3C**, **Supplementary Figure 6**). Charged residues facing the MYH7 interface had the opposite pattern where many low abundance variants are predicted to be tolerated by predictors (**Figure 3C**, **Supplementary Figure 7A**). Our results are consistent with the resolved myosin filament structure[37] and the expectation that mutating charged MYBPC3 interface residues will disrupt binding with MYH7. This discrepancy is likely due to predictors lacking contextual awareness for the binding interfaces for structural proteins like MYBPC3.

### Cell context-specific *MYBPC3* variant effect measurements have high clinical utility

Although pathogenic variants in *MYBPC3* are the leading cause of familial HCM [20,21], clinical classification of missense variants remains challenging and as a result there are currently 1,670 VUS in ClinVar. Classifying *MYBPC3* variants is difficult because many commonly used sources of evidence are not useful. For example, since HCM is common, affecting about 1 in 500 adults [38], an individual diagnosed with HCM produces little evidence towards variant pathogenicity (https://cspec.genome.network/cspec/ui/svi/doc/GN095). Since pathogenic variants have variable penetrance [39,40], family studies can be uninformative. Moreover, variant effect predictions are unreliable at interaction interfaces (**Figure 3C**). The iPSC-SGE abundance scores separate pathogenic and benign variants with near perfect accuracy, suggesting that these scores could provide much needed evidence for *MYBPC3* variant classification.

To determine how much evidence our *MYBPC3* iPSC-SGE abundance scores could provide for variant classification, we calibrated the data using known pathogenic and benign variants [41]. Our abundance scores correctly identified PLP variants as low abundance and BLB as functionally normal, equating to strong evidence (Oddspath = 58.5) for low abundance variants toward pathogenic classifications and strong evidence (Oddspath = 0.04) for functionally normal variants towards benign classifications (**Supplementary Table 1**).

We applied calibrated functional evidence to 43 VUS in the *MYBPC3* C10 domain from two clinical sources to assess the proportion of VUS that can potentially be resolved with functional data. Of these, 25 VUS were low abundance, 16 were normal, and 2 were indeterminate (**Figure 3D, Supplementary Table 2**). The majority of the low abundance VUS were located in buried residues that are also predicted to disrupt the Ig-like domain fold. In contrast, the normal abundance VUS largely occurred at exposed residues with no low abundance variants (**Supplementary Figure 8A, B**). Addition of our functional evidence would enable the reclassification of 67% of the 43 VUS, 13 to likely pathogenic, and 16 to benign or likely benign (**Figure 3E and Supplementary Table 2**). The remaining 14 VUS either had indeterminate abundance scores (14%) or lacked sufficient additional evidence beyond the functional data (86%).

Use of functional data generated in the correct cell context was a key enabler of our ability to reclassify VUS. For example, an individual evaluated at the University of Washington Adult Genetics Clinic was found to have an inherited R1205W variant and classic HCM with an abnormally thick interventricular septal wall (**Supplementary Figure 8C).** However, R1205W could not be classified as likely pathogenic due to conflicting variant effect predictions (**Supplementary Figure 5**), preventing at-risk family members from undergoing genetic testing and follow-up screening. Indeed, R1205 is a positively charged residue in contact with a negatively charged patch of the MYH7 LMM where we demonstrated that predictors perform poorly[37] (**Supplementary Figure 4C**). R1205W was low abundance in our assay, and addition of strong pathogenic evidence overcame the conflicting predictor evidence. Without our cell-type-correct abundance data, this variant and others like it would likely remain VUS, preventing patients and their families from being appropriately counseled about their HCM risk.

### Variant effect measurements in two genetic contexts reveal loss of function and dominant negative variants in *POLG*

*POLG* is a nuclear gene that encodes the DNA polymerase required for replicating the mitochondrial genome[42]. Pathogenic *POLG* variants cause inherited mitochondrial depletion syndromes, resulting in a range of neurological disorders with differing symptoms and severity. Moreover, pathogenic *POLG* missense variants have been associated with both dominant and recessive disease, making them difficult to test using current multiplexed assays[43]. Because *POLG* is essential in human pluripotent stem cells[44], we measured the effect of 437 variants in exon 16 on cellular fitness in the background of a *POLG* null and partial function allele. (**Figure 4A**). The *POLG* iPSC variant libraries were harvested 14 days after sorting for the fluorescent tags marking correctly edited variant libraries and variants were scored for their effect on fitness by comparing their abundance after the outgrowth to the starting plasmid library. We used the distribution of synonymous variants to classify variants as either depleted or functionally normal.

**Figure 4:**
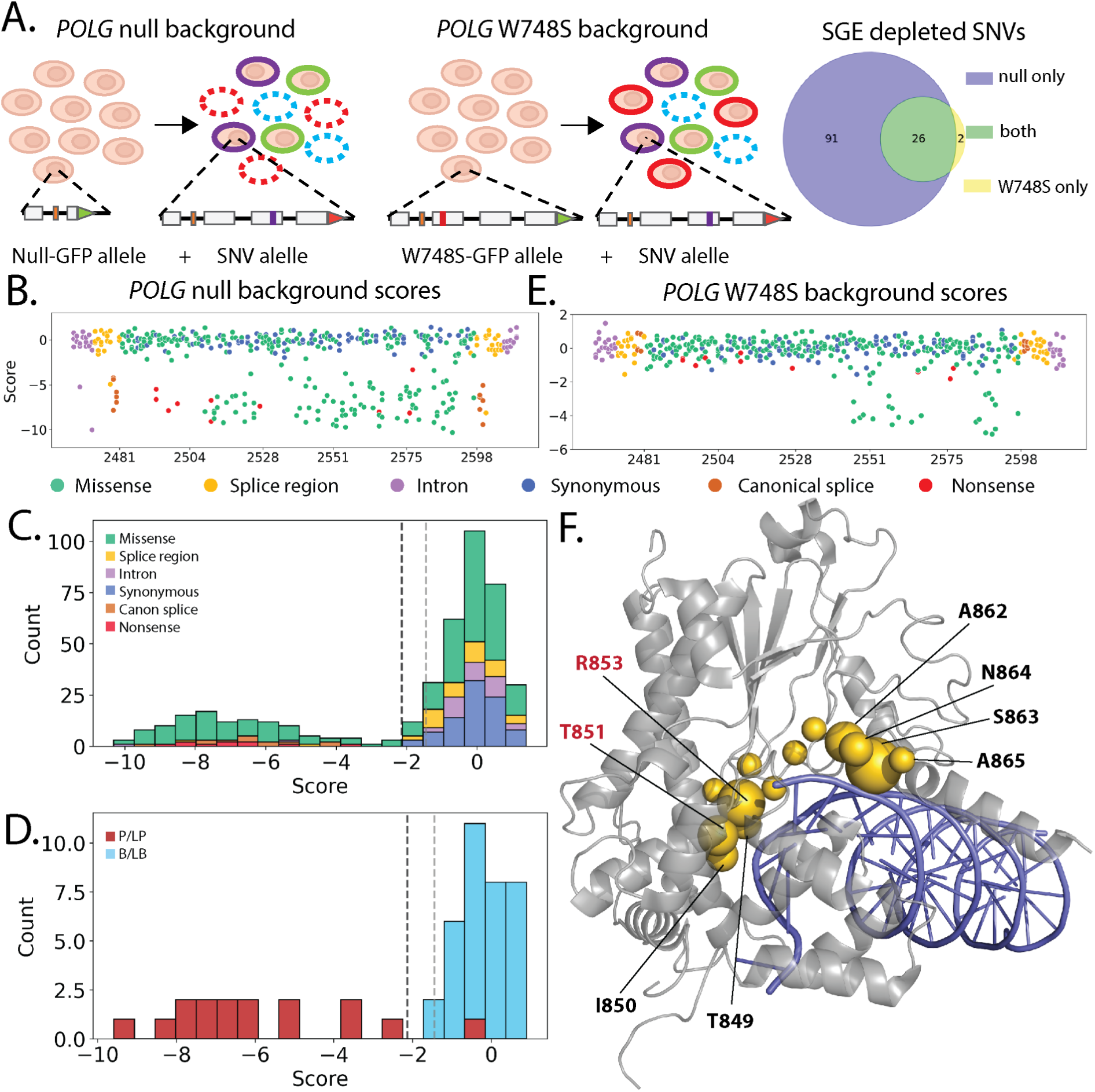
Measuring the effect of *POLG* variants on cellular fitness in iPSC. **A**) Schematic showing the background alleles for the *POLG* saturation genome editing. The exons 16 SNV library was edited in the null background (left) and partial function W748S allele (right). Loss of function variants become depleted from the edited population. A Venn diagram shows the number SNVs depleted in the null vs. W748S background. **B**) Fitness scores for *POLG* variants by genomic coordinate in the null genetic background with SNVs colored by molecular consequence. **C**) Histogram of all *POLG* fitness scores in the null genetic background colored by molecular consequence. Vertical dotted lines represent the thresholds for the functional classes. **D**) Histogram of *POLG* fitness scores for ClinVar variants colored by classification. **E**) Fitness scores for *POLG* variants by genomic coordinate in the W748S background with SNVs colored by molecular consequence. **F**) Structure showing the POLG polymerase domain bound to DNA (pdb_00008d33). The polymerase domain backbone is gray and DNA is blue and dominant negative positions have gold spheres. The number of depleted SNVs at each amino acid in the W748S background is depicted by the size of the sphere at the beta-carbon. Residue labels are red at positions with dominant fitness effects in yeast [47].

In cells with a null *POLG* background allele, all nonsense and canonical splice site variants were depleted along with 95 missense and 4 intronic variants (**Figure 4B, C; Supplementary Table 3**). All 34 BLB variants were functionally normal and 15 of 16 PLP were depleted (**Figure 4D**). With the exception of two splice region variants, all iPSC-SGE depleted variants had deleterious scores across the five predictors (**Supplementary Figure 9**).

In cells with a partial-function W748S background *POLG* allele, only 23 variants were depleted, all of which were missense. These variants occurred at 12 residues spanning amino acid positions 847 to 866 (**Figure 4E**), which make up the junction of the polymerase thumb and palm subdomains. These residues are involved in binding of the polymerase domain to template and nascent DNA strands[45]. Disruption of contacts made by two of these residues, T851 and R853, with DNA causes dominant cell growth defects in diploid yeast, suggesting that these residues are critical to mitochondrial health even in the context of a functioning second allele[46,47]. Further, additional dominant variants have been described in the polymerase domain[48], leading to the hypothesis that disrupted DNA binding leads to replication fork stalling, DNA damage, and depletion of mitochondrial DNA[46]. These variants occur primarily at arginine, serine, threonine, and asparagine residues known to interact with charged phosphate groups of the DNA backbone and are in close contact to both the template and newly synthesized DNA strands[49,50] (**Figure 4F**). Thus, we conclude that the 23 depleted missense variants act as dominant negatives on the partial-function W748S background.

Three of the *POLG* dominant negatives in our assay have been reported in individuals with adult onset phenotypes (T851A, R853W, and N864S) [51–53]. Our data suggests that heterozygous carriers of these dominant negatives may be at increased risk of developing adult onset *POLG* related disorders and would likely benefit from neurology evaluation. While all of the dominant negatives were depleted in the null background experiment, the addition of dominant negative evidence enables more effective genetic counseling and risk assessment for carriers.

### Generalizing iPSC-SGE to hundreds of targets

iPSC-SGE enables saturation scale variant effect measurement in iPSCs and their derivative cell types. To enable the broad application of iPSC-SGE to many genes we used CRISPR screens to identify multiplexible, cell fitness phenotypes in iPSCs and iPSC derived neurons, developed an engineering-free iPSC-SGE workflow that eliminates the need to create iPSC lines with defined background alleles, and demonstrated gene-sized repair template knock in.

To discover disease associated genes amenable to iPSC-SGE with a cellular fitness phenotype, we performed a CRISPR knock-out screen targeting 4,502 clinically-relevant genes in WTC-11 Ngn2 iPSCs, which can be readily differentiated into neuron-like cells. Our target gene list included genes associated with disease in the GenCC database[11], genes suspected to be associated with neuronal disease from genetic testing panels or, as controls, genes known to be essential in neurons[54,55]. We focused on neurological disease genes since they represent the largest number of VUS in ClinVar that are not accessible by HAP1 SGE (**Figure 1B**). Each gene was targeted by two validated gRNAs[56] (**Supplementary Table 4**), which were transduced at low MOI and sequenced after three weeks in iPSC culture (**Figure 5A**). We identified 592 significantly depleted and 21 significantly enriched genes in iPSCs (**Figure 5B, Supplementary Table 5**). Of the depleted genes which were also assessed in a HAP1 essentiality screen[57], 224 were essential only to iPSCs and 367 were essential in both cell lines (**Figure 5C**). Since all the genes in this screen are disease associated, these iPSC essential genes represent 224 new clinically relevant targets for SGE that are likely to have a cell fitness phenotype in iPSCs. In general, HAP1 cells were more reliant on oxidative phosphorylation and iPSCs were more reliant on processes regulating gene expression and protein translation (**Supplementary Figure 9A**).

**Figure 5:**
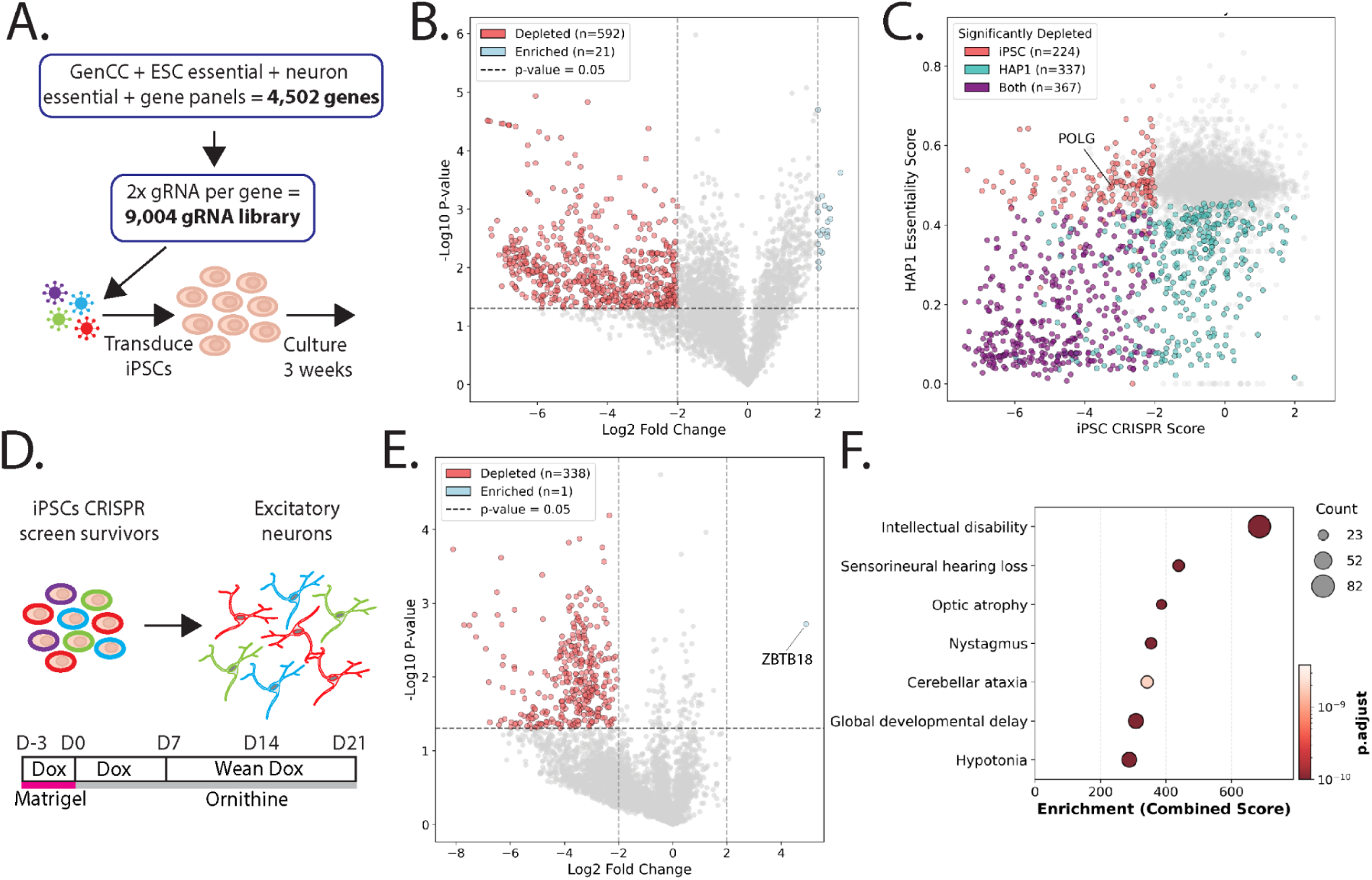
A CRISPR screen identifies disease associated genes that are essential in iPSC and iPSC-derived neurons. **A**) Schematic of CRISPR screen for iPSC cellular fitness. iPSC (WTC-11-Ngn2-Cas9) are transduced with a library of guide RNAs targeting 4,502 genes. After selection for guide integration with blasticidin, iPSCs are cultured for 3 weeks and guides are sequenced and scored vs. sgRNA plasmid library. **B)** Volcano plot of gene log2 fold change in day 21 iPSC (WTC-11-Ngn2-Cas9) vs. -log10 p-values. Significantly depleted or enriched genes are colored pink and blue respectively. **C)** Comparison of knock out screen effect sizes for genes in iPSC vs. HAP1[57], points colored as indicated. **D)** Schematic of iPSC-derived neuron CRISPR screen. Surviving cells from the WTC-11-Ngn2-Cas9 screen (A-B) were induced to differentiate into neurons [63]. Neuron differentiation is carried out until day 21 and guides were sequenced and scored from surviving cells. **E)** Volcano plot gene fold change in day 21 after neuron induction vs. iPSCs vs. -log10 P values, colors and thresholds as in B. **F)** Gene set enrichment highlighting the most significantly enriched phenotypes using Enrichr combined score [64] in neuron essential genes.

To identify disease-associated genes essential for neuronal differentiation or viability, we differentiated the transduced iPSCs into neuron-like cells after iPSC essential genes had dropped out of the population (**Figure 5D**). We identified 338 significantly depleted and one significantly enriched gene (*ZBTB18*) (**Figure 5E**). The neuron essential genes were enriched for known neuronal disorders, demonstrating the applicability of neuron viability to assess variants associated with neuronal conditions (**Figure 5F**). In total, our CRISPR screen identified 443 genes specifically essential to iPSC and/or iPSC-derived neuron survival, identifying gene targets for iPSC-SGE with a total of 95,350 current VUS.

To both validate our CRISPR screen results and highlight a potential dual editing strategy, we edited a pathogenic missense variant into *ZBTB18*, an enriched hit gene from the essentiality screen in neurons, to simultaneously make homozygous and heterozygous iPSCs. Our *MYBPC3* and *POLG* iPSC-SGE assays employed a sequential editing strategy where we engineered cell lines with desired background alleles then performed iPSC-SGE on the second allele. Generating background allele cell lines represents a workflow bottleneck, so we targeted both the background and variant alleles simultaneously in a single editing step. We generated dual editing vectors by constructing repair templates bearing either wild type *ZBTB18* or with the pathogenic R464H variant and fluorescent and drug selectable markers. We simultaneously knocked in either wild type/R464H or R464H/R464H and selected for double knock-in cells (**Figure 6A**). 19.5% of the *ZBTB18* edited cells had both alleles knocked in and were sorted to yield a population of correctly edited cells (**Figure 6B**). Our one step editing strategy dramatically decreases the time required, yielding results within 28 days as opposed to several months for the clonal background allele workflow (**Figure 6C**).

**Figure 6:**
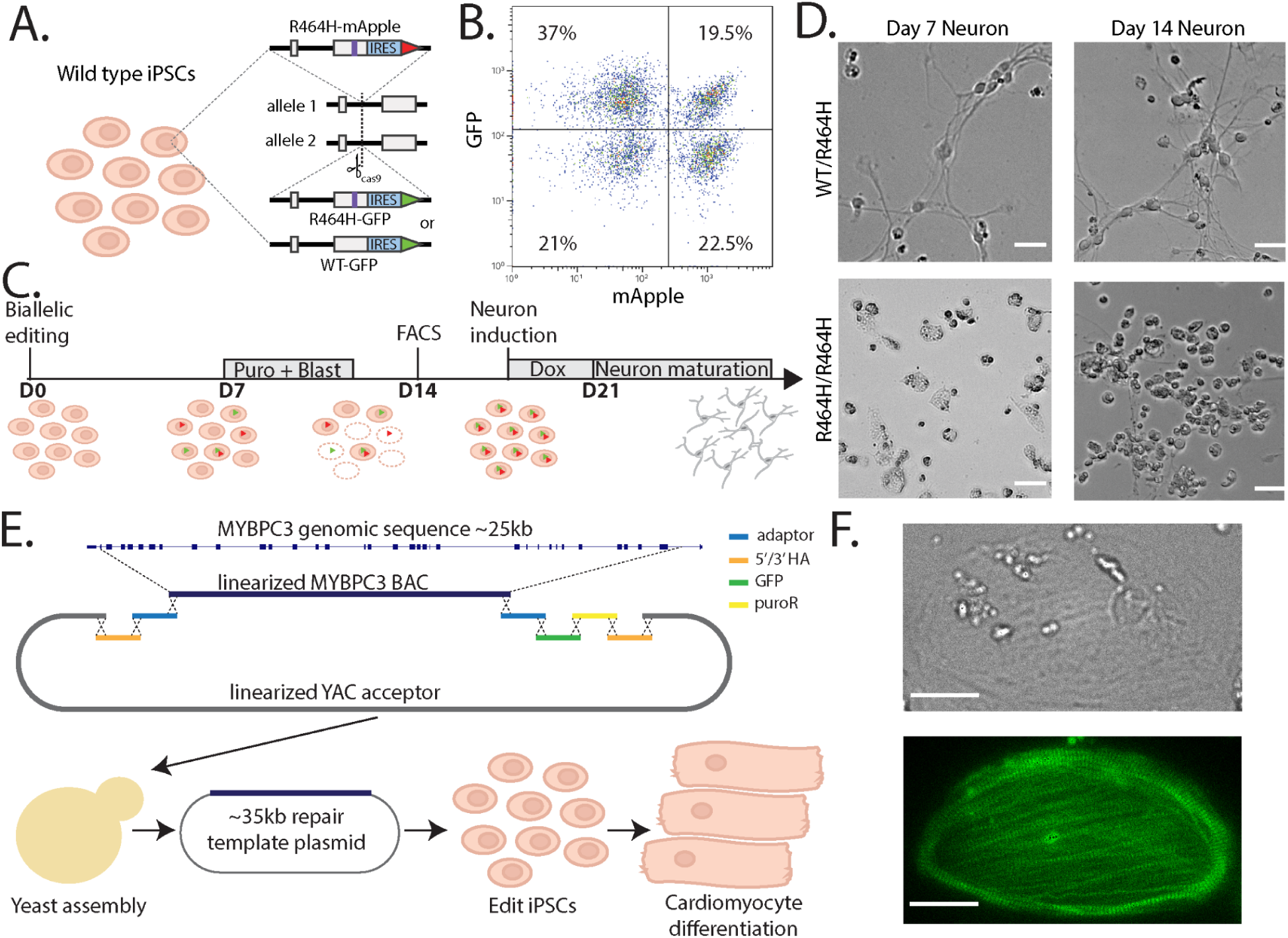
Approaches to generalize and scale iPSC-SGE. **A)** Schematic of dual editing strategy to simultaneously edit iPSC (WTC-11-Ngn2-Cas9) with *ZBTB18* wild-type(WT)/R464H or R464H/R464H alleles. **B)** FACS plot of GFP vs. mApple intensity of dual-edited iPSC. **C)** Timeline of selection for *ZBTB18* edited cells and neuronal induction. **D)** Brightfield photos of iPSC edited with *ZBTB18* wild-type(WT)/R464H or R464H/R464H alleles at 7 and 14 post induction. Scale bar indicates 20 μm. **E)** Schematic of the strategy for cloning the full length *MYBPC3* gene into a bigDNA construct followed by iPSC editing and cardiomyocyte differentiation. **F)** Brightfield (top) and fluorescent (bottom) image of big DNA-edited cardiomyocyte expressing MYBPC3. Scale bar indicates 25 μm.

Neither heterozygous nor homozygous *ZBTB18* R464H iPSC lines exhibited any obvious growth or morphological phenotypes. However, upon neuronal differentiation, homozygous R464H cells had larger cell bodies and fewer cell projections than heterozygous cells (**Figure 6D**). Additionally, the R464H homozygous cells continued to divide throughout differentiation whereas the heterozygous cell stopped dividing during maturation (**Figure 6D**). The inappropriate cell division validates the CRISPR screen result and represents a cell fitness phenotype for *ZBTB18* iPSC-SGE.

iPSC-SGE requires assembly of large repair templates. Standard cloning workflows limit the size of these repair templates, which must include introns and the selectable markers required for iPSC-SGE, to ∼15 kb of sequence. This confined our assays on *MYBPC3* and *POLG* to the 3’ end of these genes. We overcame this limitation using the yeast recombineering “big DNA” workflow [58,59] to enable gene-scale repair templates for iPSC-SGE. We assembled *MYBPC3* big DNA repair templates spanning the entire genomic sequence from exon 2 to exon 34 using a bacterial artificial chromosome (BAC) containing all of *MYBPC3* and synthesized DNA fragments containing homology arms and selectable markers needed for HDR and selection of correct edits (**Figure 6E**). We edited a reference MYBPC3-GFP big DNA repair template into WTC-11 cells, followed by differentiation into cardiomyocytes. The cardiomyocytes expressed the edited MYBPC3-GFP with green signal localized to sarcomeres, demonstrating on-target editing and correct expression pattern (**Figure 6F**).

## Discussion

Multiplexed assays of variant effect are revolutionizing precision medicine by generating functional evidence for variants that can illuminate the biology of genes, proteins and cells, reveal variant pathomechanism, and reduce the uncertainty limiting clinical genetics. However, the majority of MAVEs so far have been conducted in utilitarian cancer derived cell lines that do not account for cell or genetic context. Moreover, most human disease genes have phenotypes that are best evaluated in the relevant differentiated cell types. To address these limitations, we developed iPSC-SGE, an HDR based method for editing variants into iPSCs and measuring their effects in differentiated cell types. We developed an approach that enables efficient editing of iPSCs and used it to introduce 1,574 variants in *MYBPC3* and *POLG* on different genetic backgrounds, an ∼14-fold increase compared to previous approaches. We phenotyped these variants using several approaches, demonstrating that cell and genetic context is critical. Testing *MYBPC3* variants in cardiomyocytes enabled us to measure sarcomere-dependent variant effects and provides evidence for clinical labs to resolve 67% of VUS. Testing *POLG* variants on both null and partial function genetic backgrounds revealed both LOF variants and dominant negative variants. We also used a CRISPR screen to identify 443 potential iPSC-SGE target genes with growth phenotypes in iPSCs or iPSC derived neurons, demonstrating the generalizability of iPSC-SGE. We illustrated its scalability by substantially increasing the efficiency of biallelic editing and by enabling whole gene replacement with big DNA technology. Thus, iPSC-SGE overcomes several key barriers to the broad application of MAVEs by enabling assays in specialized cell types and multiple genetic contexts.

The editing approach that underlies iPSC-SGE improves upon other HDR-based methods by dramatically increasing scale, accommodating genetic markers that enable downstream assays and allows facile identification of on-target edits, while also preserving introns so splice effects can be measured. iPSC base editor approaches have also been developed, including for *MYBPC3 [15]*. However, base editors target both alleles and install diverse variants across the editing window, making variant effects challenging to interpret and prone to noise. Our approach allows for precise control over SNV integration, ensuring introduction of a single SNV in the targeted allele while maintaining the specified background allele.

iPSC-SGE has several limitations. First, although we demonstrated high efficiency editing across three genes, editing rates were locus specific and saturation genome editing may be challenging at other loci. However, we note that *MYBPC3* is silenced and likely heterochromatinized in iPSCs, and yet we achieved saturation editing for multiple exons suggesting broad applicability. Furthermore, we demonstrated the feasibility of editing using a ∼35kb *MYBPC3* big DNA repair template at that locus. Genes that are larger than *MYBPC3* may pose a more significant challenge to repair template synthesis and HDR efficiency, which may require further gene-specific optimizations. While our editing approach simplifies the editing workflow by using the same backbone repair template for a given gene, each exon library is introduced in an individual experiment and performing iPSC-SGE for an entire gene may require many experiments. Finally, endogenous genome editing helps maintain endogenous expression levels of variants, but the fluorescent markers and antibiotic resistant cassettes introduced at the 3’ end of the repair templates could influence gene expression levels.

By enabling MAVEs in specialized cell types and specific genetic contexts iPSC-SGE will transform how variant effects are measured. For example, massively parallel reporter assays that assess the effect of noncoding variants using transcriptional reporter assays do not always faithfully recapitulate variant effects in the absence of surrounding genomic sequence [60]. iPSC-SGE preserves noncoding sequences, meaning that noncoding variant effects can be measured in context. In addition, MAVEs have been performed on single genetic backgrounds, but iPSC-SGE, especially in combination with gene-scale repair templates allows exploration of variant effects on multiple backgrounds including interaction between coding and noncoding variants. For example, MAVEs of coding genes could be performed in the presence of common noncoding variants from risk and protective haplotypes identified by GWAS. More broadly, iPSC-SGE overcomes the cell and genetic context hurdles, enabling MAVEs across diverse cell types with programmed genetic backgrounds.

---

**Supplementary Figure 1:**
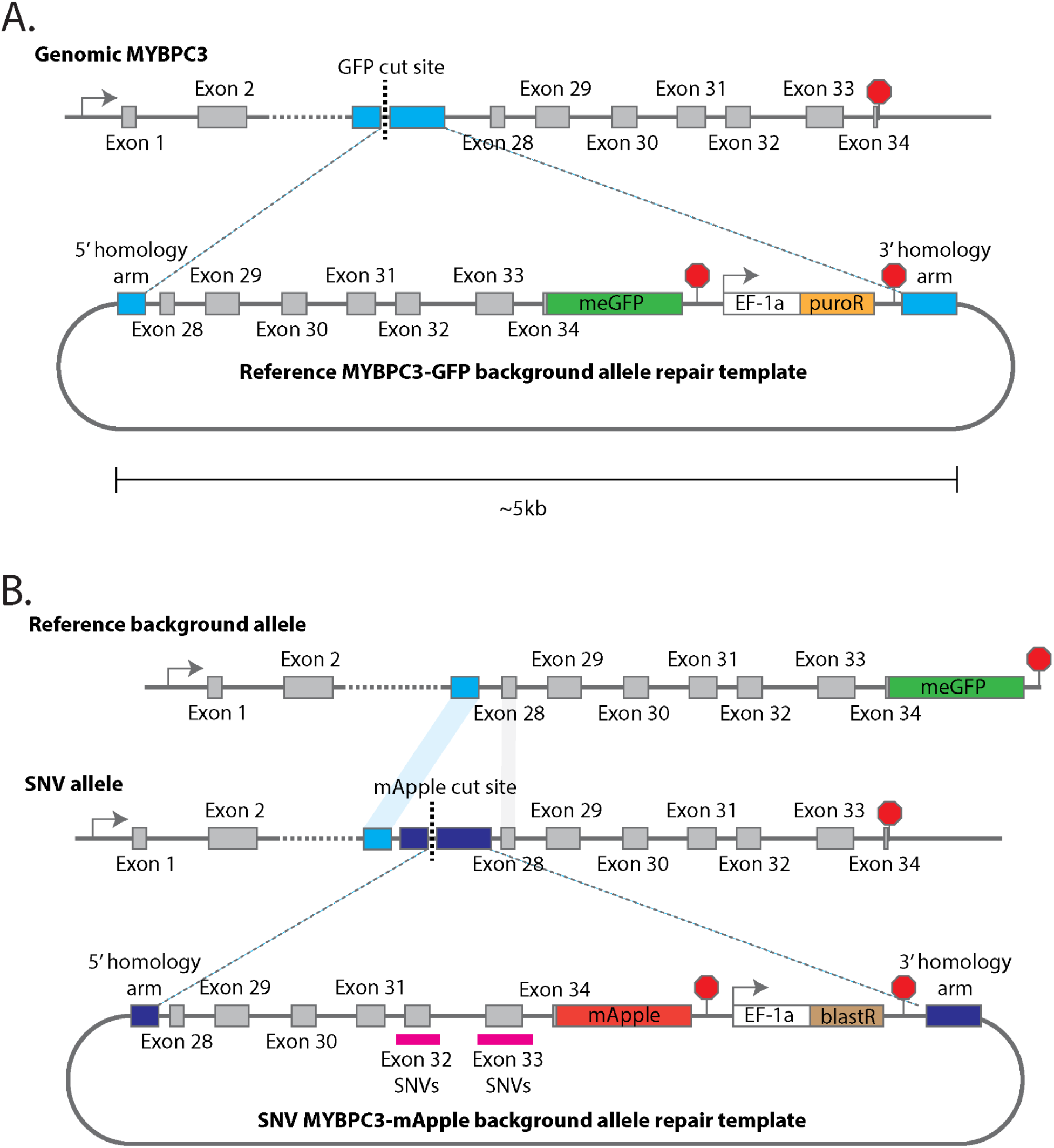
MYBPC3 iPSC-SGE repair templates. **A**) Reference background allele including meGFP fusion and puromyosin resistance cassette. Repair template has the 3’ homology arm deleted from the intronic sequence adjacent to exon 28. **B**) SNV allele repair template backbone that targets the deleted region from the background allele repair template that is still present in the wild type allele. The 3’ homology arm is also deleted from the intronic sequence adjacent to exon 28 to ensure HDR with the full repair template. SNVs were introduced into exon 32 or 33 and flanking intronic sequences.

**Supplementary Figure 2:**
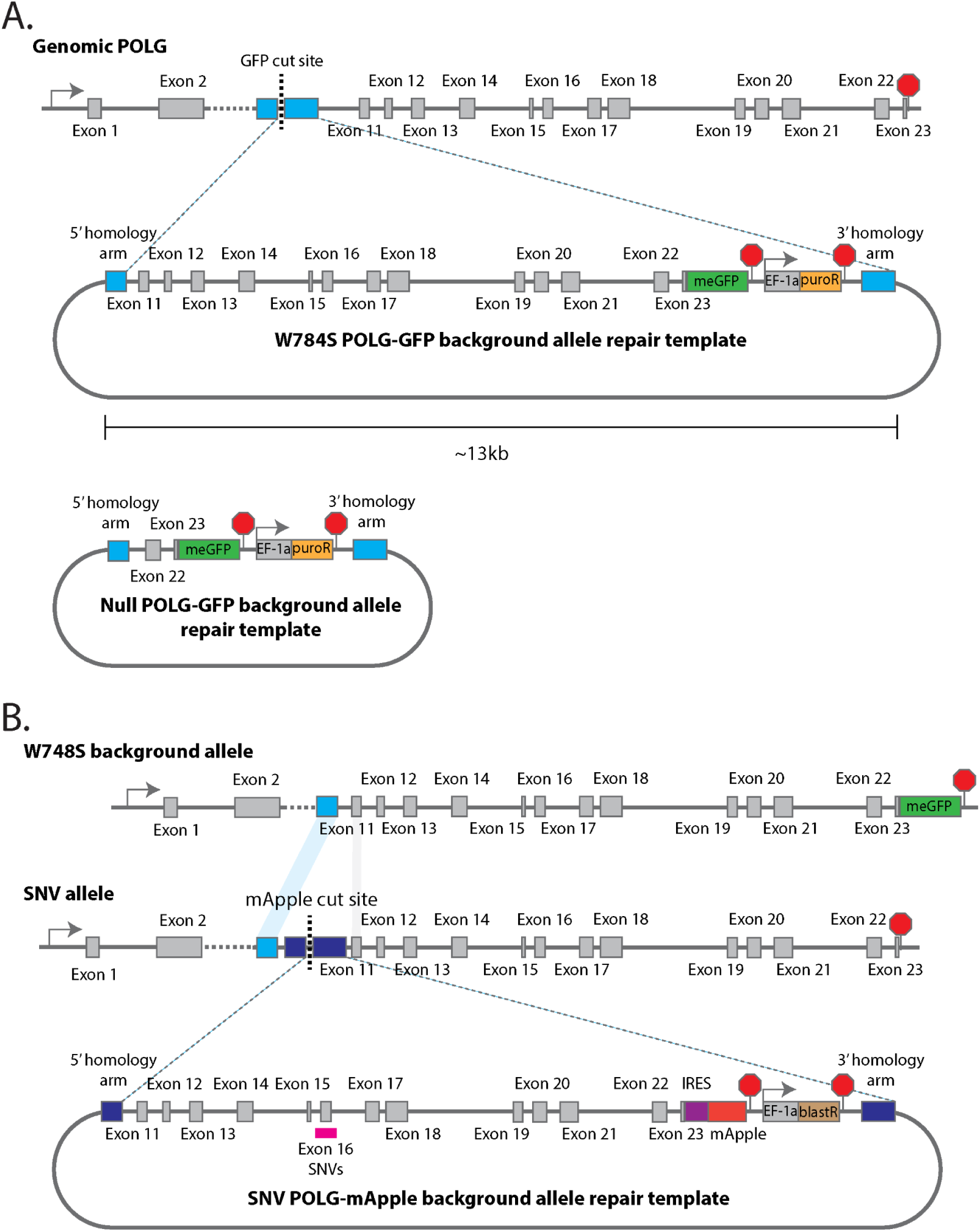
POLG iPSC-SGE repair templates: **A**) Reference background allele including meGFP fusion and puromyosin resistance cassette. W748S repair template has the 3’ homology arm deleted from the intronic sequence adjacent to exon 11. Null repair template deletes exons 11-22 and fuses meGFP to in frame deletion product. **B**) SNV allele repair template backbone that targets the deleted region from the background allele repair template that is still present in the wild type allele. The 3’ homology arm is also deleted from the intronic sequence adjacent to exon 11 to ensure HDR with the full repair template. SNVs were introduced into exon 16 and flanking intronic sequences.

**Supplementary Figure 3:**
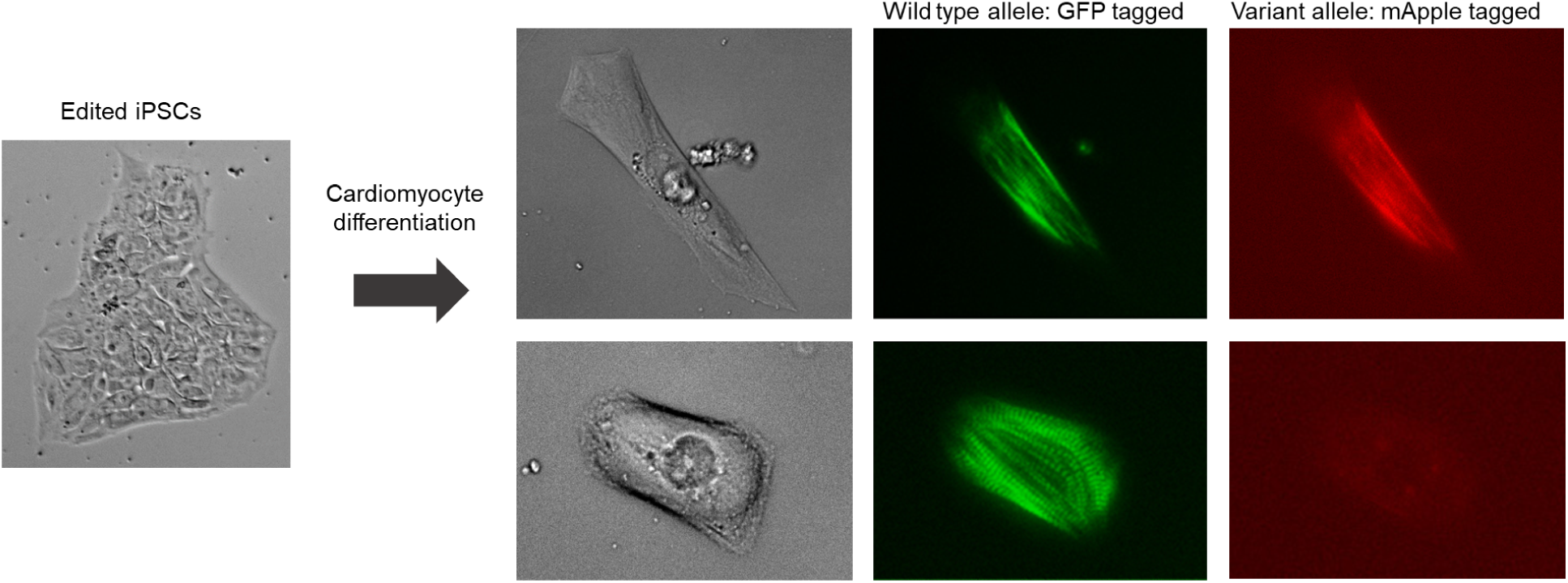
MYBPC3 iPSC-SGE cells. iPSC colony imaged after blasticidin selection for SNV allele integration. Cells were then differentiated to cardiomyocytes and dissociated and plated sparsely for imaging at differentiation day 20. Top row shows a representative wild type like cell with GFP and mApple signal localizing to sarcomeres. Bottom row shows a reduced abundance variant with normal GFP signal but very dim and diffuse mApple signal. Scale bar indicates 25 μm.

**Supplementary Figure 4:**
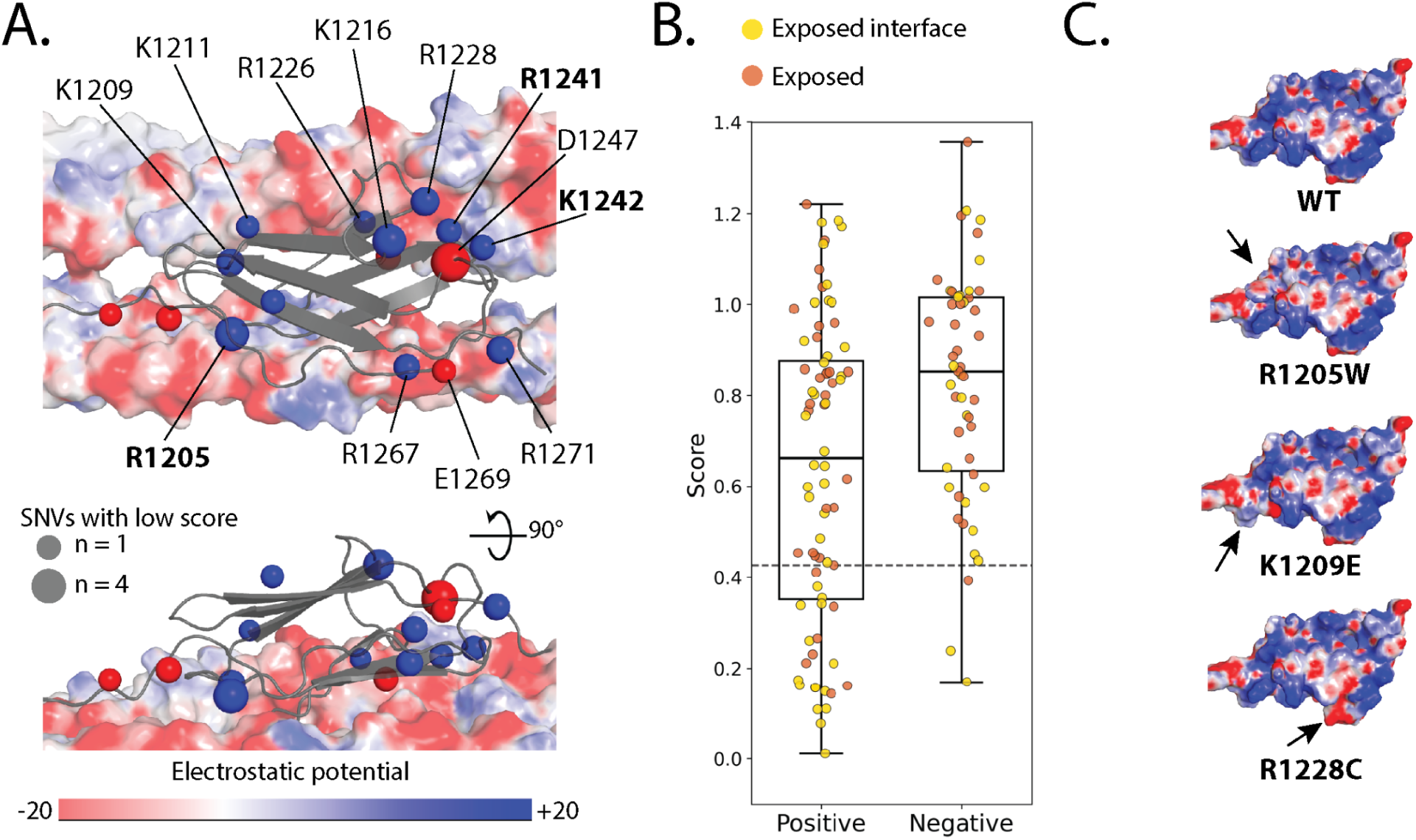
Disruption of MYBPC3 electrostatic potential causes reduced abundance. **A**) MYBPC3 C10 domain (gray backbone) with spheres representing charged residue beta carbons. The size of the sphere indicates the number of reduced abundance variants at that position. Reference negative charged residues are red and reference positive charged residues are blue. MYH7 tails are represented as a surface colored by electrostatic potential. B) Box plots representing variant scores are charged residues. Horizontal dashed line represents reduced abundance threshold. Yellow dots are exposed at the MYH7 interface and orange dots are exposed but not at the MYH7 interface. **C**) Exposed residue variants impacting electrostatic potential of C10 domain.

**Supplementary Figure 5:**
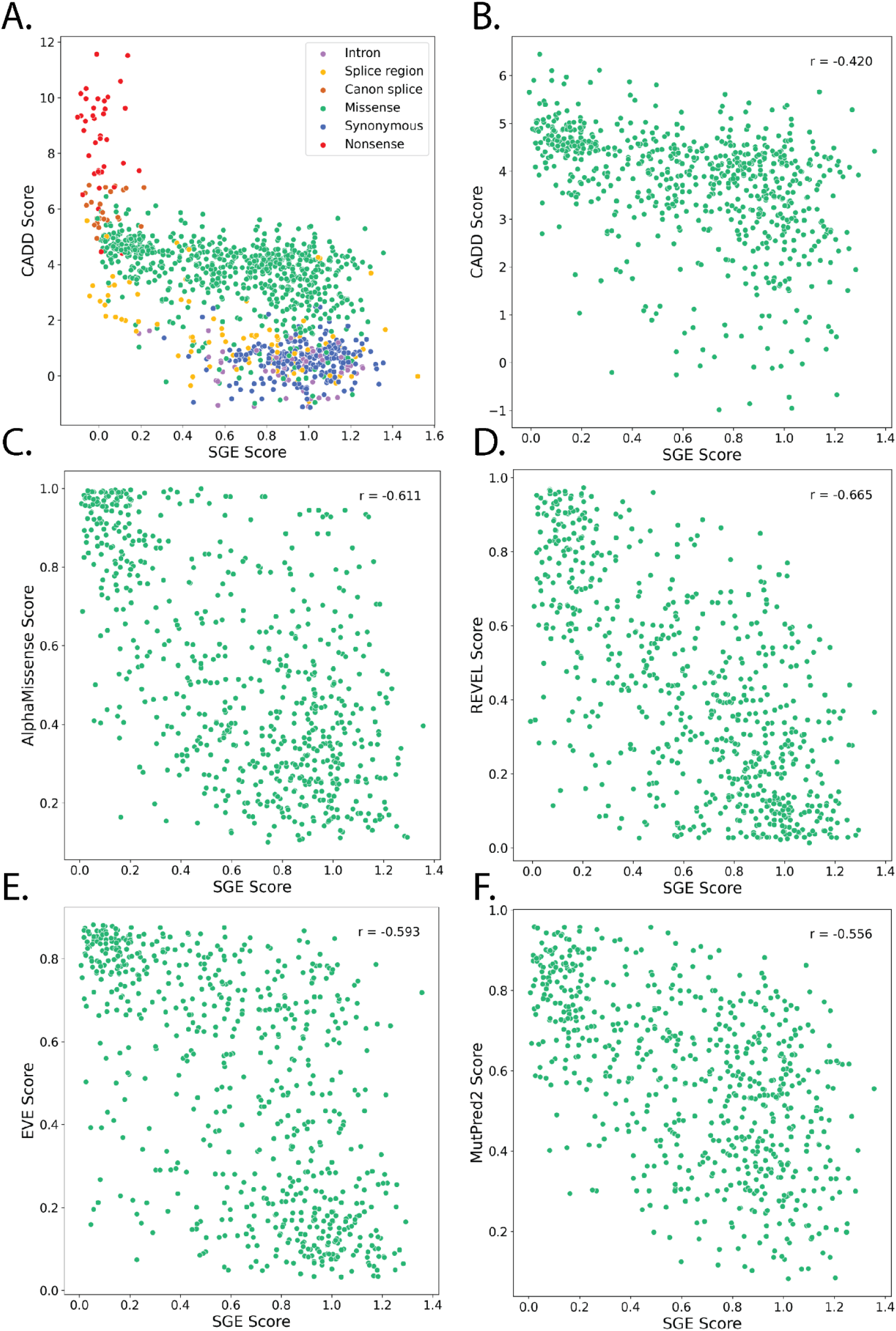
MYBPC3 iPSC-SGE correlation with variant effect predictors. Abundance scores are plotted agains VEP scores from CADD, AlphaMissense, REVEL, EVE, and MutPred2 [61,65–68]. Pearson’s r is indicated in each plot.

**Supplementary Figure 6:**
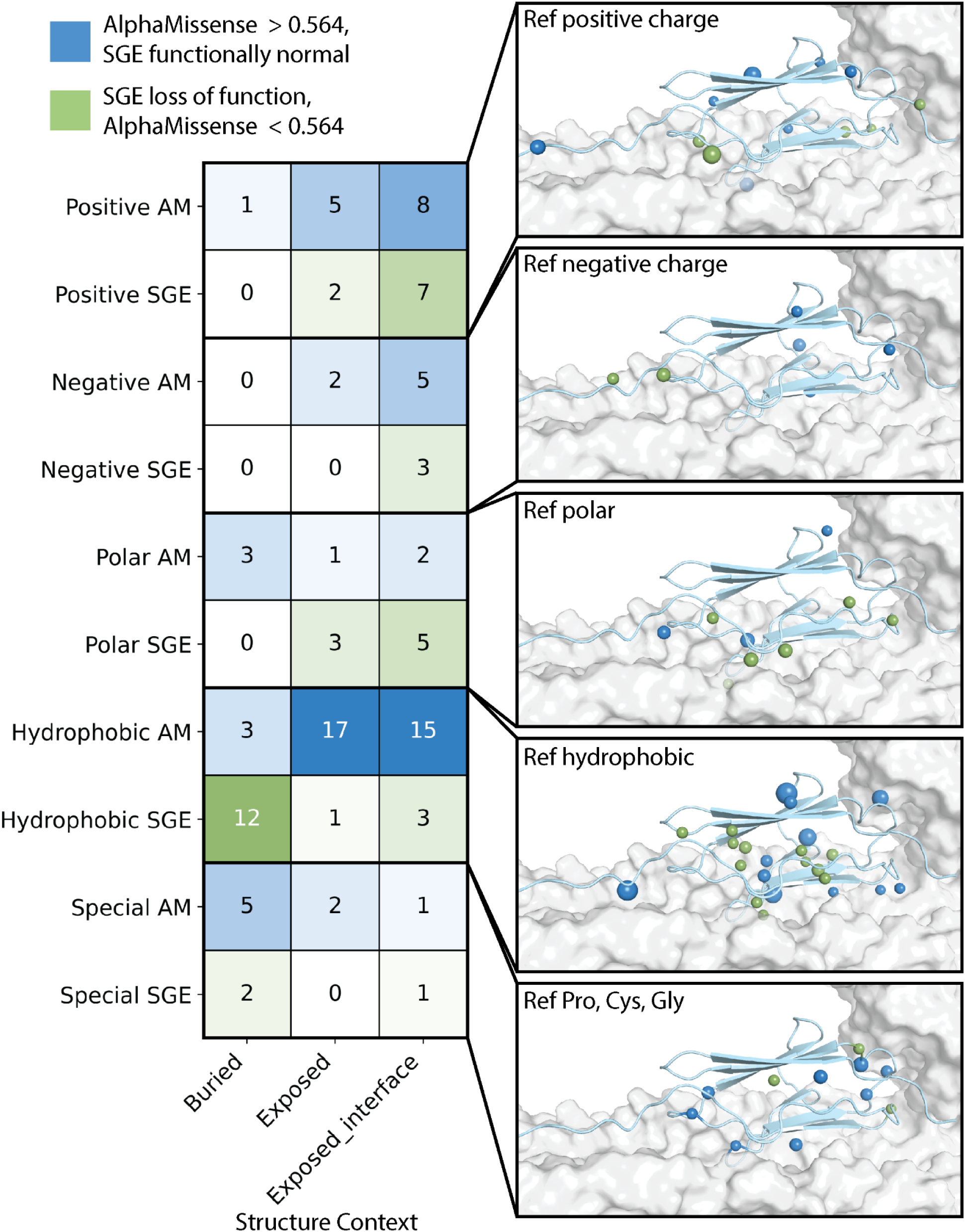
Positional discordance patterns between MYBPC3 abundance scores and AlphaMissense. Blue boxes represent variants with AlphaMissense scores above 0.564 (predicted deleterious) and normal abundance scores. Green boxes represent variants with Alphamissense scores below 0.564 and low abundance scores. Heatmap is separated by reference amino acid class as in Figure 3. Each reference group is projected individually on the MYBPC3 structure with MYH7 LMM surface. Spheres represent beta-carbons at the discordant amino acid positions, where sphere color represents the direction of discordance, and sphere size indicates the number of discordant variants at that position.

**Supplementary Figure 7:**
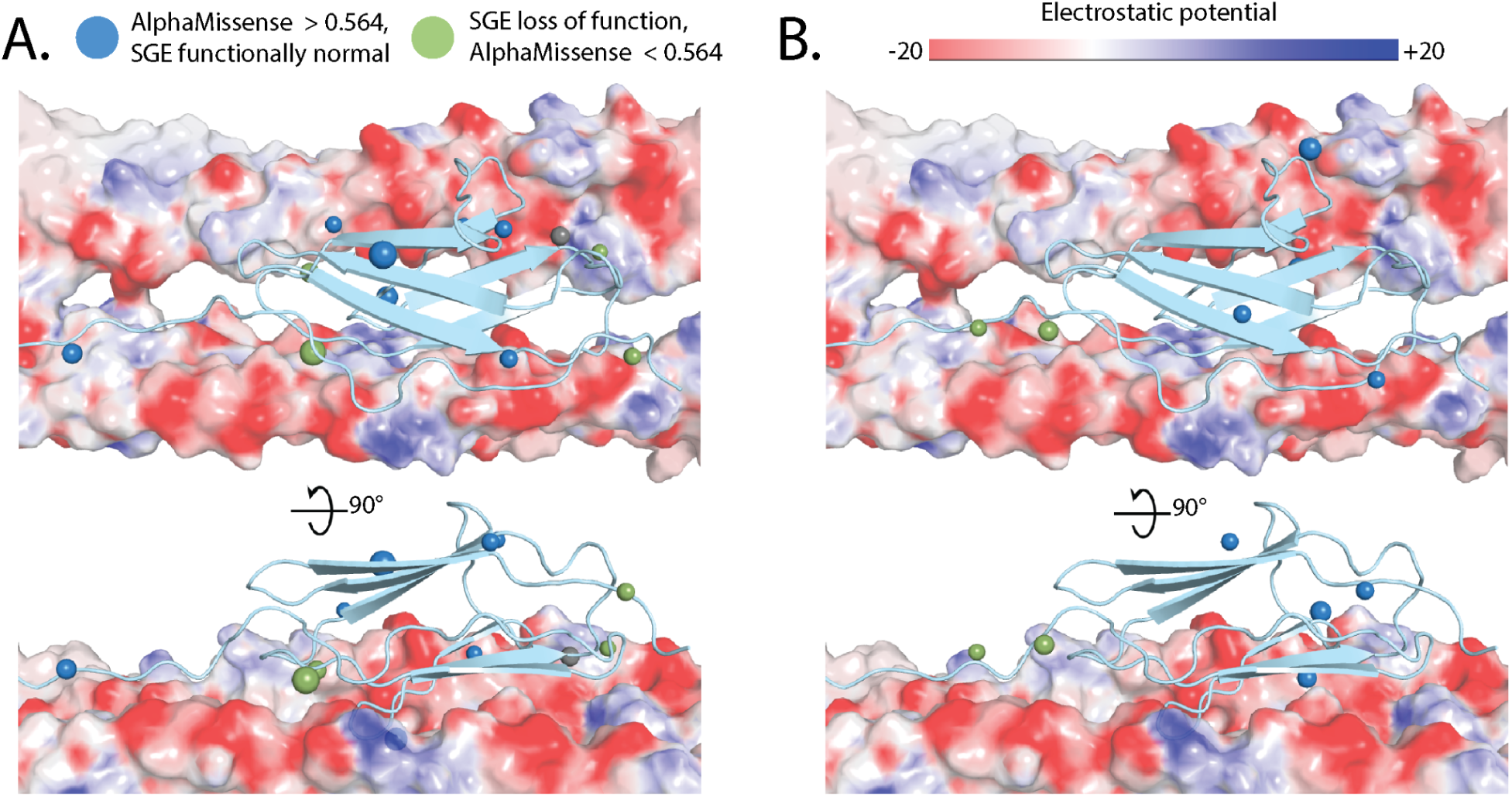
Charged residue discordance between MYBPC3 abundance and AlphaMissense. **A**) Reference positive amino acids with discordant scores represented with spheres. Blue spheres represent variants with AlphaMissense scores above 0.564 (predicted deleterious) and normal abundance scores. Green spheres represent variants with Alphamissense scores below 0.564 and low abundance scores. MYH7 LMM surface electrostatic potential is represented where red is more negative and blue is more positive. **B**) Reference negative amino acid discordant positions represented as in A.

**Supplementary Figure 8:**
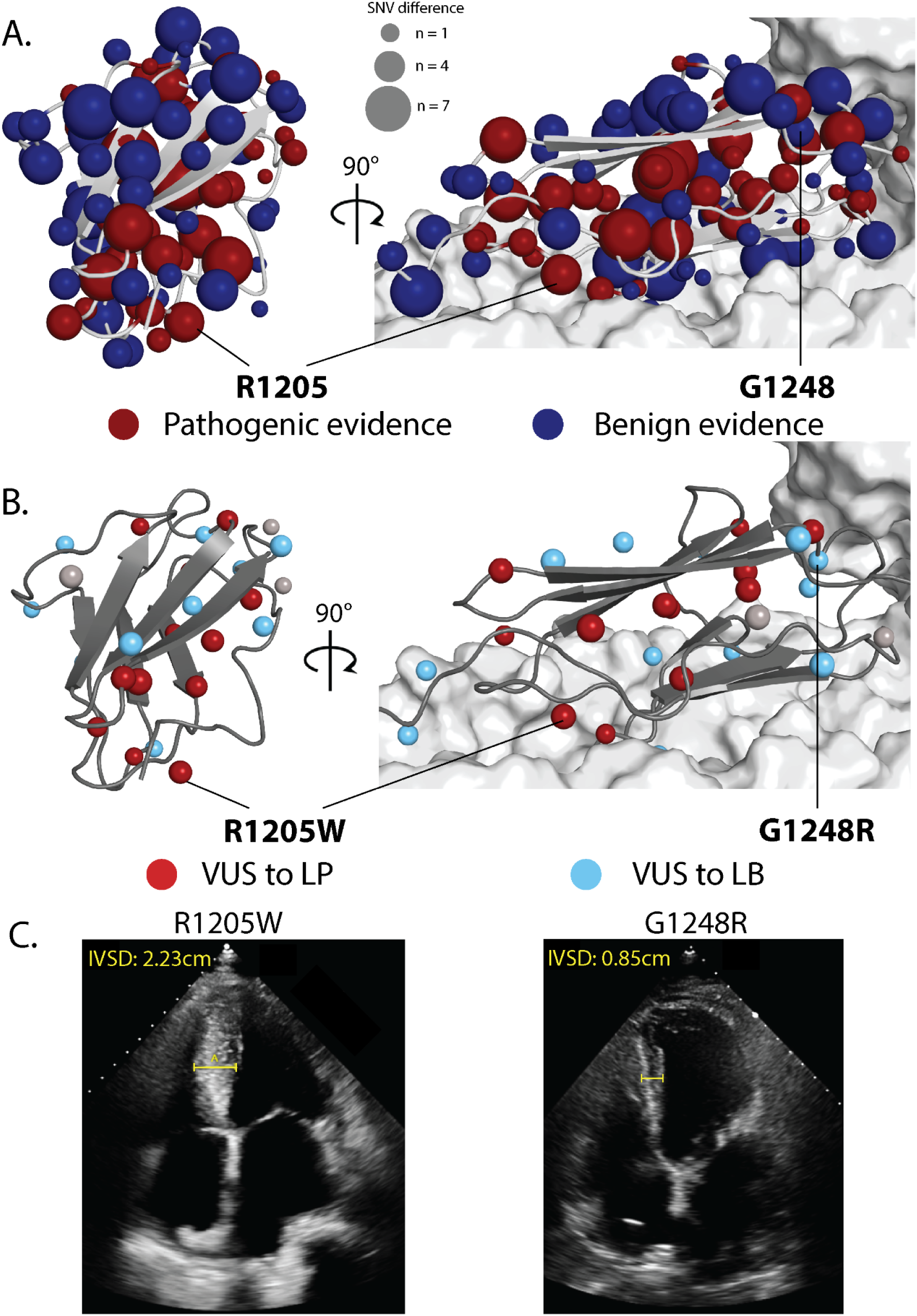
VUS reclassification follows positional abundance patterns. **A**) MYBPC3 abundance scores represented on MYBPC3 structure with MYH7 LMM surface. Spheres represent beta-carbons for assayed positions. Red spheres represent positions with more low abundance scores than normal abundance and blue spheres represent positions with more normal abundance scores than low abundance. Sphere size represents the difference. **B**) VUS reclassifications represented on the structure. Light blue represents VUS to LB, red represents VUS to LP, and gray represents VUS to VUS. Size of the sphere corresponds to reclassified VUS count. **C)** Echocardiograms of UWMC patients with indicated *MYBPC3* variants

**Supplementary Figure 9:**
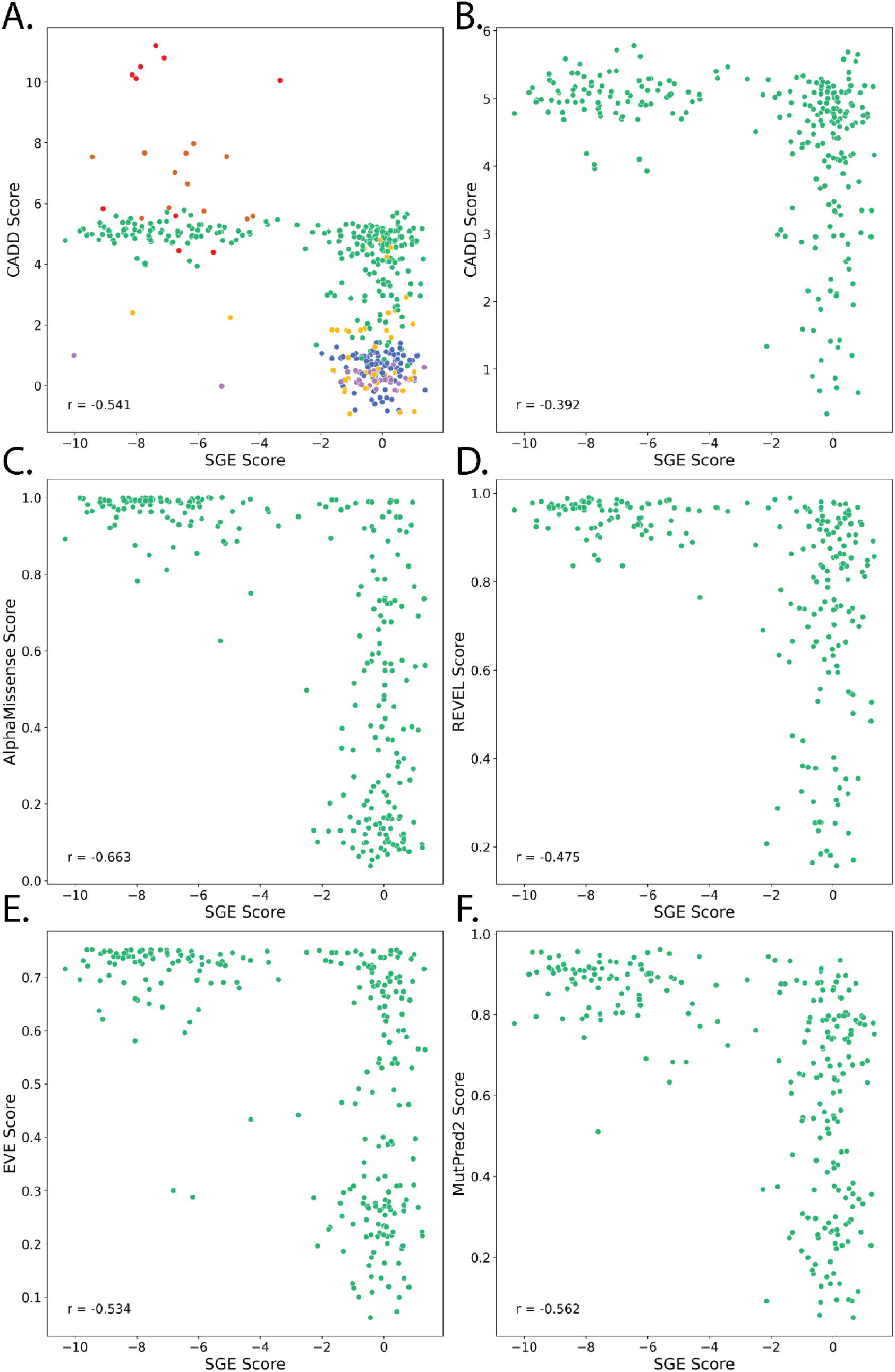
POLGiPSC-SGE correlation with variant effect predictors. POLG fitness scores are plotted against VEP scores from CADD, AlphaMissense, REVEL, EVE, and MutPred2 [61,65–68]. Pearson’s r is indicated in each plot.

## Resource Availability

### Lead contacts

Further information and requests for resources and reagents should be directed to and will be fulfilled by the lead contacts, Shawn Fayer, Douglas M. Fowler and Lea M. Starita.

### Materials availability

Please contact the corresponding author with requests for any reagents generated by this study.

### Data and code availability

Code to reproduce data analysis and generating figures can be found at: https://github.com/sfayer31/iPSC-SGE-manuscript

## Acknowledgements

We thank members of the Starita, Fowler, and Yang labs for helpful discussion. This work was funded by the NIH NHGRI Centers of Excellence in Genome Sciences (5RM1HG010461) and the Brotman Baty Institute Catalytic Collaboration (BBI2019CC17). R.K.G. acknowledges support from the Washington Research Foundation postdoctoral fellowship. The Brotman Baty Institute Clinical Variant Database (BBI-CVD) is funded by a Brotman Baty Institute Catalytic Collaboration (BBI2021CC10) and the National Human Genome Research Institute (R01HG013025) to A.B.S. and L.M.S.. C.A.G. is the Akiko Yamazaki and Jerry Yang Faculty Scholar in Pediatric Translational Medicine, Stanford Maternal & Child Health Research Institute.

## Ethics statement

Use of de-identified human sequence data from Ambry Genetics was approved by University of Washington IRB study #3598 and the use of identifiable human sequence and phenotype data from the Brotman Baty Institute Clinical Variant Database by UW IRB study #14771. MEK and EDS are paid employees of Ambry Genetics.

## Methods

### iPSC culture and maintenance

Human induced pluripotent stem cells (iPSCs) were maintained at 37°C in 5% CO2. All WTC-11, WTC-11-Ngn2 [63] and derivative lines generated for this project were cultured on Matrigel (Corning; cat. no. 354277) and fed every other day with mTeSR Plus and 0.5% penicillin-streptomycin (ThermoFisher; cat. no. 15140122). iPSCs were passaged before reaching confluency by dissociating cells with TrypLE Express (ThermoFisher; cat. no. 12604013) and resuspended in mTeSR Plus supplemented with 10 μM Y-27632 dihydrochloride (Rho kinase [ROCK] inhibitor; Tocris; cat. no. 1254). Clonal lines were isolated via dilution plating in 96 well plates at 0.5 cells per well in mTeSR Plus supplemented with 10 μM Y-27632 and CloneR2 (StemCell Technologies; cat. no. 100-0691). iPSCs were transferred from Matrigel to plates coated with Vitronectin XF (StemCell Technologies; cat. no.07180) at 10 µg/ml and conditioned for at least two passages before cardiac differentiation on Vitronectin.

### Construction of iPSC-SGE repair templates

Plasmid repair templates were assembled using Twist gene fragments containing the genomic region of interest, homology arms, fluorescent protein fusion, and antibiotic selection markers driven by Ef1ɑ. Homology arms flanked the genomic Cas9 cut site and were between 200-500 bases in length. Gene fragments were designed with 18-22 base overlaps and assembled together with assembly PCR. Assembled repair templates were then cloned into the pJET vector with directional cloning. SNVs were inserted into mApple fused repair templates with either Gibson assembly or Golden Gate assembly. MYBPC3 exons 32 and 33 SNV libraries (**Supplementary Figure 1**) were amplified from Twist oligo pools and inserted into the linearized backbone with Gibson assembly with NEBuilder HiFi DNA assembly (NEB; cat. no. E2621S). The *POLG* backbone vector was over 13 kb and could not be linearized for Gibson assembly and instead the Twist fragment containing exon 16 was designed with type IIS restriction sites and SNV oligos were amplified with compatible cut sites (**Supplementary Figure 2**). SNVs were then inserted using Golden Gate assembly. All repair templates were sequence verified with whole plasmid sequencing via long read sequencing technologies before integration into cells. Assemblies were electroporated into NEB 10-beta electrocompetent cells (NEB; cat. no. C3020K) and plasmids were prepped with the ZymoPURE II Plasmid Maxipre Kit (Zymo; cat. no. D4203).

### Big DNA

*MYBPC3* big DNA repair templates were assembled from a bacterial artificial chromosome (BAC) containing the full *MYBPC3* genomic sequence. The BAC was linearized for assembly with synthesized gene fragments containing the 5’ and 3’ homology arms, meGFP, and puromycin resistance cassette. Fragments contained ∼100bp overlap and were transformed and assembled in yeast as previously described [59]. Sequence correct clones are then transformed into *E. coli* and prepped with the ZymoPURE II Plasmid Maxiprep Kit (Zymo; cat. no. D4203). The big DNA repair template targets the first intron, maintaining the endogenous promoter and first exon so expression is dependent on correct integration into the *MYPBC3* locus in iPSCs.

### gRNA design

Guides were designed for *POLG* intron 10 and *MYBPC3* intron 27. Guides were optimized for predicted cutting efficiency and minimal predicted off-target effects with ccTOP (https://cctop.cos.uni-heidelberg.de/). The guides for inserting the background GFP allele were designed 5’ to the guide for SNV integration so that the guide sequence for the SNV allele could be removed from the GFP repair template. Synthetic guide RNAs and SpCas9 protein were ordered from Synthego (https://synthego.com).

### Generation of heterozygous GFP lines for SGE

Wild type WTC-11 iPSCs were grown to 70-80% confluency then split 1:5 the day before gene editing. Synthetic gRNAs were pre-complexed into ribonucleoproteins (RNPs) with SpCas9 protein for 10 minutes at room temperature per the manufacturer recommendations (Synthego). RNPs and 1 µg of plasmid repair template were introduced into 200,000 cells via electroporation with the Lonza Amaxa P3 Primary Cell Kit S (Lonza; cat. No. V4XP-3032) using the Lonza 4D Nucleofector system. Immediately after electroporation, cells were plated into a single well of a 24 well plate coated with Matrigel and supplemented with 1X Clone R2. One week post electroporation, cells were selected with 1 µg/ml puromycin for 2 days. Cells were then expanded and sorted for GFP positivity. Since *MYBPC3* is not expressed in stem cells, correct integration of repair templates was determined with CRaTER, which used CRISPR activation at the endogenous promoter to turn on *MYBPC3[69]* 3 days prior to the sort. GFP positive cells were then plated in 96 well plates at 0.5 cells per well and single cell colonies were selected and expanded for PCR verification of heterozygous integration of GFP repair templates.

### Saturation genome editing of iPSCs

Clonal GFP iPSCs were grown to 70-80% confluency then split 1:5 the day before gene editing. Synthetic gRNAs were pre-complexed into ribonucleoproteins (RNPs) with SpCas9 protein for 10 minutes at room temperature per the manufacturer recommendations (Synthego). RNPs and SNV containing plasmid repair template (4 µg for *POLG* and 8 µg for *MYBPC3*) were introduced into 2x10^6^ cells via electroporation with the Lonza Amaxa P3 Primary Cell Kit L (Lonza; cat. No. V4XP-3024). Immediately after electroporation, cells were plated into a single well of a 6 well plate coated with Matrigel and supplemented with 1X Clone R2 and 10 μM Y-27632. Cells were expanded and plated sparsely in Matrigel coated 10 cm dishes for selection with 10 µg/ml blasticidin S HCl (ThermoFisher; cat. no. A1113902). After selection cells were expanded and frozen in 500 µl CryoStor CS-10 (Sigma; cat. no. C2874) at 3x10^6^ cells per vial. *POLG* cells were sorted for double positive GFP/mApple signal and expanded before cryopreservation.

### Monolayer cardiomyocyte directed differentiation

iPSCs were cultured on 10 µg/ml Vitronectin XF prior to cardiomyocyte differentiation. Small molecule directed differentiation was performed as previously described with some modifications [69]. iPSCs were plated in 15 µg/ml Vitronectin XF coated 24 well plates at 7.5x10^4^ cells per well in mTeSR Plus supplemented with 10 μM Y-27632. Media was changed to fresh mTeSR Plus without 10 μM Y-27632 the next day. Directed differentiation was initiated (D0) when cells reached 60-70% confluency by aspirating mTeSR and replacing media with RBA media: RPMI (Invitrogen; cat. no. 11875135), 0.5 mg/mL bovine serum albumin (Sigma; cat. no. A9418), 0.213 mg/mL ascorbic acid (Sigma; cat. no. A8960) supplemented with 4.5 μM CHIR-99021 (TOCRIS; cat. no. 4423). After two days (D2), CHIR-99021 containing media was removed and replaced with RBA supplemented with 2 μM Wnt-C59 (Selleck; cat. no. S7037). On D4, Wnt-C59-containing media was replaced with RBA. On D6 (and every other day afterwards), media was replaced with cardiomyocyte media: RPMI, B27 plus insulin (Invitrogen; cat. no. 17504044). Cardiomyocyte cultures typically begin beating by D6. After D12, cardiomyocyte media was removed and cardiomyocytes were dissociated to single cells with TrypLE Select 10X (ThermoFisher; cat. no. A1217701) and replated on Matrigel coated 6 well plates at 3x10^6^ cells per well in cardiomyocyte media supplemented with 10% FBS and 10 μM Y-27632.

### Monolayer cardiomyocyte and sorting

After D20 and at least 7 days after replating cardiomyocytes were dissociated to single cells with TrypLE Select 1X (ThermoFisher; cat. no. 12563011) and recovered in RPMI. Cells were washed with 1X PBS and fixed with 4% paraformaldehyde (ThermoFisher; cat. no. 043368.9M) for 10 minutes at room temperature and washed with 1X PBS. Cardiomyocytes were then resuspended in PBS sort buffer containing 5mM EDTA (cat. no. 15575020) and 25mM HEPES (ThermoFisher; cat. no.15630080). Resuspended cells were passed through a 100uM cell strainer into FACS tubes. GFP positive cells were then sorted into 4 quartile bins on mApple to GFP ratio. After the sort, cells were pelleted in Eppendorf tubes at 800 RCF for 10 minutes and supernatant was discarded.

### Monolayer cardiomyocyte DNA extraction

Cardiomyocyte DNA was extracted from sorted cardiomyocytes using the MagMAX Multi Sample Ultra 2.0 Kits (ThermoFisher; cat. no.A36570) with the MagMAX Cell and Tissue Extraction DNA Extraction Buffer (ThermoFisher; cat. No. A45469). DNA was extracted to manufacturers specifications with a few modifications. First, cell pellets of no more than 1x10^6^ cells were resuspended in extraction buffer and heated to 65C with shaking for 60 minutes with vortexing every 10 minutes. Further, the lysis step on the instrument was performed for 15 minutes. Finally, DNA was eluted from beads in 100 µl of water.

### Cardioid differentiation

Cardioid differentiation protocol was adapted from [70] [71] as previously described in [28] with some modifications. iPSCs were cultured on 5 μg/mL Vitronectin (ThermoFisher; cat. no. A14700) in mTeSR Plus(StemCell Technologies; cat. no. 100-0276) until reaching 70-80% confluency. Cells were then dissociated with Accutase (StemCell Technologies; cat. no. 07920), seeded into 96-well round-bottom ultra-low attachment plates (Corning; cat. no. 7007) in mTeSR Plus supplemented with 10 μM Y-27632 (Tocris; cat. no. 1254) at a density of 10,000 cells per well, and spun down at 900 RPM for 3 minutes at 25°C. Differentiation was induced the next day (D0) by changing the media to FlyAB(Ins), consisting of RPMI (ThermoFisher; cat. no. 11875119), B-27 supplement (ThermoFisher; cat. no. 17504044), 10 ng/mL BMP4 (Fisher; cat. no. 314BP050), 30 ng/mL FGF2 (Fisher; cat. no. 23-3FB-500CF), 5 μM LY294002 (Tocris; cat. no. 1130), 4 μM CHIR 99021 (Tocris; cat. no. 4423), and 4 ng/mL Activin A (R&D Systems; cat. no. 338-AC). On D1, media was replenished with fresh FlyAB(Ins). At 36-40 hours post-induction (D1.5), media was changed to BWIIFRa, consisting of RPMI, B-27 supplement, 10 ng/mL BMP4, 8 ng/mL FGF2, 5 μM IWP 2 (Tocris; cat. no. 3533), 0.5 μM retinoic acid (Tocris; cat. no. 0695), and 200 ng/mL VEGF (R&D Systems; cat. no. 293-VE). On D2.5-4.5, media was replenished with fresh BWIIFRa. On D5.5, media was changed to BFI, consisting of RPMI, B-27 supplement, 10 ng/mL BMP4, 8 ng/mL FGF2, and 200 ng/mL VEGF. On D6.5, media was replenished with fresh BFI. On D7.5, media was changed to maintenance media, consisting of RPMI and B-27 supplement. Maintenance media was replenished every 48 hours.

### Cardioid immunofluorescence and microscopy

Cardioid immunofluorescence and microscopy was performed as previously described [28] with slight modifications. Cardioids were fixed in 4% paraformaldehyde (PFA) and embedded in Optimal Cutting Temperature (OCT) compound (Tissue-Tek, 4583). Samples were cryosectioned at 10 µm using a cryomicrotome (Thermo Scientific; Microm HM 550), and adjacent sections were collected on coated glass slides (VWR, VWRU48311-703). Sections from the central region of each organoid were selected for analysis. For immunofluorescence staining, sections were fixed again in 4% PFA for 20 minutes and washed with PBS. Samples were blocked with 5% goat serum (BioAbChem, 72-0480) in PBS containing 0.1% Triton X-100 (MP Biochemicals, 194854) for 1 hour at room temperature, then incubated overnight at 4°C with primary antibodies. After three PBS washes, sections were incubated with fluorescently conjugated secondary antibodies for 2 hours at room temperature and washed again. Finally, slides were mounted with DAPI Fluoromount-G (Southern Biotech, 0100-20) and imaged using a Zeiss LSM 990 confocal microscope.

### Cardioid dissociation and sorting

After D18, cardioids were dissociated into single cells for fluorescence-activated cell sorting (FACS). Cardioids were harvested and washed in PBS on a bi-directional 37 μm filter (StemCell Technologies; cat. no. 27215), then dissociated using TrypLE Select 10X (Gibco; cat. no. A1217701) in a thermomixer set to 900 RPM and 37°C for 10 minutes with gentle pipetting every 5 minutes. Dissociated cardioids were pelleted at 200 x g for 3 minutes, resuspended in PBS + 1% BSA (Sigma; A1595), and passed through 35 μm cell strainer (Fisher; cat. no. 08-771-23). Cells were sorted using a Sony MA900 and 100 μm sorting chip (Sony; cat. no. LE-C3210) into the following bins: 1) GFP-/tdTomato-, 2) GFP+/tdTomato-, 3) GFP+/tdTomato+, and 4) GFP-/tdTomato+. Following FACS, cells were collected at 200 x g for 3 minutes, then resuspended in Bambanker (Fisher, cat. no. 50-999-554) for cryopreservation.

### Variant classification

We calibrated our *MYBPC3* abundance scores for clinical classification following the current ClinGen-approved calibration method to generate Bayesian likelihood ratios (OddsPath) supporting PS3/BS3 criteria [41]. We used the synonymous distribution to define normal abundance, indeterminate, and low abundance classes where z-score above -2 was considered normal abundance, z-score less than -3 was low abundance and between -2 and -3 was indeterminate. Using ClinVar P/LP and B/LB variants with at least 1 star, we calculated the OddsPath for the normal abundance (0.038) and low abundance (58.5) score intervals (**Supplementary Table 1**), which correspond to BS3 and PS3 strong evidence, respectively. Clinically observed SNVs from *MYBPC3* exons 32 and 33 and flanking intronic regions were obtained from Ambry Genetics and the Brotman Baty Institute Clinical Variant Database. We collected a total of 43 VUS from these clinical sources and followed *MYBPC3* VCEP guidance with several adaptations to reclassify VUS. For VEP data we used REVEL with updated ClinGen evidence strength thresholds [72]. We also removed PM1 hotspot evidence since this is potentially circular with VEP evidence. Additionally, we did not consider PM2 allele frequency evidence for pathogenic interpretation. (**Supplementary Table 2**).

### *POLG* cell fitness assay

After sorting genome edited *POLG* cells for positive expression of the background GFP allele and SNV mApple allele, cells were cultured for an additional 14 days. After 14 days, iPSCs were collected and genomic DNA was extracted using the Qiagen DNeasy Blood and Tissue Kit (Qiagen; cat. no. 69504).

### Illumina library preparation and sequencing

Sequencing libraries were prepared with a three step nested PCR approach. First, SNV allele specific PCR was performed using primer sets that amplified from mApple to a genomic region outside of the 5’ homology arm of the SNV repair template. *MYBPC3* allele specific PCR was conducted with Q5 Hot Start High-Fidelity 2x Master Mix (NEB; cat. no. M0494S). *POLG* allele specific PCR was performed with LongAmp Taq DNA Polymerase (NEB; cat. no.M0323S). gDNA PCRs were split into 4 reactions with 250 ng gDNA each and pooled for AMPure XP Bead (Beckman Coulter; cat. no. A63882) cleanup. Exon specific amplicons with Illumina TruSeq and Nextera adaptors were prepared in a second PCR that was AMPure cleaned. The third PCR attached ten base pair barcodes to amplicons for demultiplexing. 2 nM libraries were pooled and sequenced on the Illumina Nextseq 2000 instrument.

### Calculating iPSC-SGE scores for *MYBPC3* abundance assay

Abundance scores were calculated as previously described [23]. Briefly, sequence reads were filtered to include only SNVs mapping to the genomic regions of *MYBPC3* exon 32 or 33. Weighted average frequencies were calculated by the sum of weighted frequencies for a given variant across all bins divided by the sum of frequencies for that variant across all bins. Bins were weighted as follows: bin 1 (lowest abundance) = 0.1, bin 2 = 0.15, bin 3 = 0.2, and bin 4 (highest abundance) = 1.

### Calculating iPSC-SGE scores for *POLG* cell fitness assay

SNV log2 ratios were calculated by variant frequency at iPSC day 14 vs. pDNA frequency. First, reads were demultiplexed using Illumina Bcl2Fastq 2.20 software and paired end reads were assembled using Pear v0.9.11 software with parameters: -n 30, -m 300, -q 30. Any reads with more than one SNV or an indel were filtered out of the analysis. Reads with a single SNV were mapped to the genomic region of *POLG* exon 16 and each SNV was assigned a count. Log2 ratios were calculated for each experiment relative to plasmid DNA frequencies.

### CRISPR screen guide cloning

gRNA sequences for the 4,502 human disease genes in our CRISPR screen were from the minimal Cas9 library including the 200 non-targeting controls (**Supplementary Table 3**)[56]. Guides were ordered as an oligo pool from Twist and cloned into LentiGuide Blast (Addgene:199622) with Golden Gate assembly. Three replicates of NEB 10-Beta electrocompetent cells were transformed with the assembled gRNA library and plasmids were prepped with ZymoPURE II Plasmid Maxiprep Kit. Plasmids were pooled and sequenced with full plasmid sequencing to verify correct assembly and amplicon sequencing of the gRNA region to verify library coverage for lentivirus production.

### iPSC and induced neuron CRISPR screens

WTC-11-Ngn2 iPSCs[73] were engineered to integrate a SpCas9 expression cassette into the genome via PiggyBac transposition. Lentivirus was produced by co-transfecting HEK-293T cells with the gRNA library containing LentiGuide-Blast plasmid along with other lentivirus packaging component plasmids. After three days, lentivirus containing supernatant was collected and lentivirus was concentrated with PEG-it (System Biosciences; cat. no. LV810A-1). Ngn2-Cas9 iPSCs were seeded into Matrigel coated 6 well plates at 2x10^5^ cells per well in mTeSR Plus supplemented with 10 μM Y-27632. The following day cells were transduced with concentrated lentivirus at low MOI (about 30% transduction efficiency). To achieve 200x coverage of the gRNA library, 6x10^6^ cells were transduced per replicate. Post transduction, cells were selected with blasticidin and expanded to maintain library complexity. Cell pellets were harvested for library screening after blasticidin selection. gRNA containing iPSCs were cultured for 3 weeks, cell pellets were collected, then cells were plated for neuron differentiation following published protocols [63]. Induced neurons were collected after 21 days of maturation. Phenotype enrichment analysis was performed using Enrichr [64] to identify phenotypes associated with neuron-essential genes. Results were ranked by Enrichr combined score, which integrates both fold enrichment and statistical significance.

## References

1. Findlay GM, Daza RM, Martin B, Zhang MD, Leith AP, Gasperini M, et al. Accurate classification of BRCA1 variants with saturation genome editing. Nature. 2018;562: 217–222.

2. Buckley M, Terwagne C, Ganner A, Cubitt L, Brewer R, Kim D-K, et al. Saturation genome editing maps the functional spectrum of pathogenic VHL alleles. Nat Genet. 2024;56: 1446–1455.

3. Funk JS, Klimovich M, Drangenstein D, Pielhoop O, Hunold P, Borowek A, et al. Deep CRISPR mutagenesis characterizes the functional diversity of TP53 mutations. Nat Genet. 2025;57: 140–153.

4. Erwood S, Bily TMI, Lequyer J, Yan J, Gulati N, Brewer RA, et al. Saturation variant interpretation using CRISPR prime editing. Nat Biotechnol. 2022;40: 885–895.

5. Sahu S, Galloux M, Southon E, Caylor D, Sullivan T, Arnaudi M, et al. Saturation genome editing-based clinical classification of BRCA2 variants. Nature. 2025;638: 538–545.

6. Radford EJ, Tan H-K, Andersson MHL, Stephenson JD, Gardner EJ, Ironfield H, et al. Saturation genome editing of DDX3X clarifies pathogenicity of germline and somatic variation. Nat Commun. 2023;14: 7702.

7. Waters AJ, Brendler-Spaeth T, Smith D, Offord V, Tan HK, Zhao Y, et al. Saturation genome editing of BAP1 functionally classifies somatic and germline variants. Nat Genet. 2024;56: 1434–1445.

8. Olvera-León R, Zhang F, Offord V, Zhao Y, Tan HK, Gupta P, et al. High-resolution functional mapping of RAD51C by saturation genome editing. Cell. 2024;187: 5719–5734.e19.

9. Huang H, Hu C, Na J, Hart SN, Gnanaolivu RD, Abozaid M, et al. Functional evaluation and clinical classification of BRCA2 variants. Nature. 2025;638: 528–537.

10. Landrum MJ, Chitipiralla S, Kaur K, Brown G, Chen C, Hart J, et al. ClinVar: updates to support classifications of both germline and somatic variants. Nucleic Acids Res. 2025;53: D1313–D1321.

11. DiStefano MT, Goehringer S, Babb L, Alkuraya FS, Amberger J, Amin M, et al. The Gene Curation Coalition: A global effort to harmonize gene-disease evidence resources. Genet Med. 2022;24: 1732–1742.

12. Friedman CE, Fayer S, Pendyala S, Chien W-M, Loiben A, Tran L, et al. CRaTER enrichment for on-target gene editing enables generation of variant libraries in hiPSCs. J Mol Cell Cardiol. 2023;179: 60–71.

13. Lv W, Qiao L, Petrenko N, Li W, Owens AT, McDermott-Roe C, et al. Functional Annotation of TNNT2 Variants of Uncertain Significance With Genome-Edited Cardiomyocytes. Circulation. 2018;138: 2852–2854.

14. Blanch-Asensio A, Grandela C, Brandão KO, de Korte T, Mei H, Ariyurek Y, et al. STRAIGHT-IN enables high-throughput targeting of large DNA payloads in human pluripotent stem cells. Cell Rep Methods. 2022;2: 100300.

15. Yamamoto Y, Chua K, Ferrasse A, Kirilova A, De Jong HN, Floyd BJ, et al. Scaled multidimensional assays of variant effect identify sequence-function relationships in hypertrophic cardiomyopathy. bioRxiv. 2025. doi:10.1101/2025.05.23.655878

16. Helms AS, Thompson AD, Glazier AA, Hafeez N, Kabani S, Rodriguez J, et al. Spatial and Functional Distribution of Pathogenic Variants and Clinical Outcomes in Patients With Hypertrophic Cardiomyopathy. Circ Genom Precis Med. 2020;13: 396–405.

17. Friedman CE, Fayer S, Pendyala S, Chien W-M, Loiben A, Tran L, et al. CRaTER enrichment for on-target gene editing enables generation of variant libraries in hiPSCs. J Mol Cell Cardiol. 2023;179: 60–71.

18. Silva AC, Moreira JN, Lobo JMS, Almeida H. Current Applications of Pharmaceutical Biotechnology. Springer Nature; 2020.

19. Rath A, Mishra A, Ferreira VD, Hu C, Omerza G, Kelly K, et al. Functional interrogation of Lynch syndrome-associated MSH2 missense variants via CRISPR-Cas9 gene editing in human embryonic stem cells. Hum Mutat. 2019;40: 2044–2056.

20. Alfares AA, Kelly MA, McDermott G, Funke BH, Lebo MS, Baxter SB, et al. Results of clinical genetic testing of 2,912 probands with hypertrophic cardiomyopathy: expanded panels offer limited additional sensitivity. Genet Med. 2015;17: 880–888.

21. Ho CY, Day SM, Ashley EA, Michels M, Pereira AC, Jacoby D, et al. Genotype and Lifetime Burden of Disease in Hypertrophic Cardiomyopathy: Insights from the Sarcomeric Human Cardiomyopathy Registry (SHaRe). Circulation. 2018;138: 1387–1398.

22. Helms AS, Thompson AD, Glazier AA, Hafeez N, Kabani S, Rodriguez J, et al. Spatial and Functional Distribution of Pathogenic Variants and Clinical Outcomes in Patients With Hypertrophic Cardiomyopathy. Circ Genom Precis Med. 2020;13: 396–405.

23. Matreyek KA, Starita LM, Stephany JJ, Martin B, Chiasson MA, Gray VE, et al. Multiplex assessment of protein variant abundance by massively parallel sequencing. Nat Genet. 2018;50: 874–882.

24. Pocock MW, Reid JD, Robinson HR, Charitakis N, Krycer JR, Foster SR, et al. Maturation of human cardiac organoids enables complex disease modeling and drug discovery. Nat Cardiovasc Res. 2025;4: 821–840.

25. Holman AR, Tran S, Destici E, Farah EN, Li T, Nelson AC, et al. Single-cell multi-modal integrative analyses highlight functional dynamic gene regulatory networks directing human cardiac development. Cell Genom. 2024;4: 100680.

26. Desai D, Song T, Singh RR, Baby A, McNamara J, Green LC, et al. D389V Variant Induces Hypercontractility in Cardiac Organoids. Cells. 2024;13. doi:10.3390/cells13221913

27. Li Y, Ma K, Dong Z, Gao S, Zhang J, Huang S, et al. Frameshift variants in C10orf71 cause dilated cardiomyopathy in human, mouse, and organoid models. J Clin Invest. 2024;134. doi:10.1172/JCI177172

28. Metzl-Raz E, Zhao R, Deshpande S, Powell J, Porter EG, Zouaghi Y, et al. Deep learning the dynamic regulatory sequence code of cardiac organoid differentiation. bioRxiv. 2025. doi:10.1101/2025.10.15.680997

29. Regalado SG, Qiu C, Lalanne J-B, Martin BK, Duran M, Trapnell C, et al. Barcoded monoclonal embryoids are a potential solution to confounding bottlenecks in mosaic organoid screens. bioRxiv. 2025. doi:10.1101/2025.05.23.655669

30. Carrier L, Bonne G, Bährend E, Yu B, Richard P, Niel F, et al. Organization and sequence of human cardiac myosin binding protein C gene (MYBPC3) and identification of mutations predicted to produce truncated proteins in familial hypertrophic cardiomyopathy. Circ Res. 1997;80: 427–434.

31. Marston S, Copeland O ’neal, Gehmlich K, Schlossarek S, Carrier L. How do MYBPC3 mutations cause hypertrophic cardiomyopathy? J Muscle Res Cell Motil. 2012;33: 75–80.

32. Previs MJ, Beck Previs S, Gulick J, Robbins J, Warshaw DM. Molecular mechanics of cardiac myosin-binding protein C in native thick filaments. Science. 2012;337: 1215–1218.

33. Previs MJ, Prosser BL, Mun JY, Previs SB, Gulick J, Lee K, et al. Myosin-binding protein C corrects an intrinsic inhomogeneity in cardiac excitation-contraction coupling. Sci Adv. 2015;1. doi:10.1126/sciadv.1400205

34. Einheber S, Fischman DA. Isolation and characterization of a cDNA clone encoding avian skeletal muscle C-protein: an intracellular member of the immunoglobulin superfamily. Proc Natl Acad Sci U S A. 1990;87: 2157–2161.

35. Okagaki T, Weber FE, Fischman DA, Vaughan KT, Mikawa T, Reinach FC. The major myosin-binding domain of skeletal muscle MyBP-C (C protein) resides in the COOH-terminal, immunoglobulin C2 motif. J Cell Biol. 1993;123: 619–626.

36. Miyamoto CA, Fischman DA, Reinach FC. The interface between MyBP-C and myosin: site-directed mutagenesis of the CX myosin-binding domain of MyBP-C. J Muscle Res Cell Motil. 1999;20: 703–715.

37. Dutta D, Nguyen V, Campbell KS, Padrón R, Craig R. Cryo-EM structure of the human cardiac myosin filament. Nature. 2023;623: 853–862.

38. Maron BJ, Gardin JM, Flack JM, Gidding SS, Kurosaki TT, Bild DE. Prevalence of hypertrophic cardiomyopathy in a general population of young adults. Echocardiographic analysis of 4111 subjects in the CARDIA Study. Coronary Artery Risk Development in (Young) Adults. Circulation. 1995;92: 785–789.

39. Topriceanu C-C, Pereira AC, Moon JC, Captur G, Ho CY. Meta-Analysis of Penetrance and Systematic Review on Transition to Disease in Genetic Hypertrophic Cardiomyopathy. Circulation. 2024;149: 107–123.

40. Lorenzini M, Norrish G, Field E, Ochoa JP, Cicerchia M, Akhtar MM, et al. Penetrance of Hypertrophic Cardiomyopathy in Sarcomere Protein Mutation Carriers. J Am Coll Cardiol. 2020;76: 550–559.

41. Brnich SE, Abou Tayoun AN, Couch FJ, Cutting GR, Greenblatt MS, Heinen CD, et al. Recommendations for application of the functional evidence PS3/BS3 criterion using the ACMG/AMP sequence variant interpretation framework. Genome Med. 2019;12: 3.

42. Hance N, Ekstrand MI, Trifunovic A. Mitochondrial DNA polymerase gamma is essential for mammalian embryogenesis. Hum Mol Genet. 2005;14: 1775–1783.

43. Rahman S, Copeland WC. POLG-related disorders and their neurological manifestations. Nat Rev Neurol. 2019;15: 40–52.

44. Yilmaz A, Peretz M, Aharony A, Sagi I, Benvenisty N. Defining essential genes for human pluripotent stem cells by CRISPR-Cas9 screening in haploid cells. Nat Cell Biol. 2018;20: 610–619.

45. Lee Y-S, Kennedy WD, Yin YW. Structural insight into processive human mitochondrial DNA synthesis and disease-related polymerase mutations. Cell. 2009;139: 312–324.

46. Stumpf JD, Copeland WC. Mitochondrial DNA replication and disease: insights from DNA polymerase γ mutations. Cell Mol Life Sci. 2011;68: 219–233.

47. Stumpf JD, Bailey CM, Spell D, Stillwagon M, Anderson KS, Copeland WC. mip1 containing mutations associated with mitochondrial disease causes mutagenesis and depletion of mtDNA in Saccharomyces cerevisiae. Hum Mol Genet. 2010;19: 2123–2133.

48. Jazayeri M, Andreyev A, Will Y, Ward M, Anderson CM, Clevenger W. Inducible expression of a dominant negative DNA polymerase-gamma depletes mitochondrial DNA and produces a rho0 phenotype. J Biol Chem. 2003;278: 9823–9830.

49. Luscombe NM, Laskowski RA, Thornton JM. Amino acid-base interactions: a three-dimensional analysis of protein-DNA interactions at an atomic level. Nucleic Acids Res. 2001;29: 2860–2874.

50. Park J, Herrmann GK, Mitchell PG, Sherman MB, Yin YW. Polγ coordinates DNA synthesis and proofreading to ensure mitochondrial genome integrity. Nat Struct Mol Biol. 2023;30: 812–823.

51. Woodbridge P, Liang C, Davis RL, Vandebona H, Sue CM. POLG mutations in Australian patients with mitochondrial disease. Intern Med J. 2013;43: 150–156.

52. Rempe T, Kuhlenbäumer G, Krüger S, Biskup S, Matschke J, Hagel C, et al. Early-onset parkinsonism due to compound heterozygous POLG mutations. Parkinsonism Relat Disord. 2016;29: 135–137.

53. Davidzon G, Greene P, Mancuso M, Klos KJ, Ahlskog JE, Hirano M, et al. Early-onset familial parkinsonism due to POLG mutations. Ann Neurol. 2006;59: 859–862.

54. Lu C, Garipler G, Dai C, Roush T, Salome-Correa J, Martin A, et al. Essential transcription factors for induced neuron differentiation. Nat Commun. 2023;14: 8362.

55. Tian R, Gachechiladze MA, Ludwig CH, Laurie MT, Hong JY, Nathaniel D, et al. CRISPR Interference-Based Platform for Multimodal Genetic Screens in Human iPSC-Derived Neurons. Neuron. 2019;104: 239–255.e12.

56. Gonçalves E, Thomas M, Behan FM, Picco G, Pacini C, Allen F, et al. Minimal genome-wide human CRISPR-Cas9 library. Genome Biol. 2021;22: 40.

57. Blomen VA, Májek P, Jae LT, Bigenzahn JW, Nieuwenhuis J, Staring J, et al. Gene essentiality and synthetic lethality in haploid human cells. Science. 2015;350: 1092–1096.

58. Brosh R, Laurent JM, Ordoñez R, Huang E, Hogan MS, Hitchcock AM, et al. A versatile platform for locus-scale genome rewriting and verification. Proc Natl Acad Sci U S A. 2021;118. doi:10.1073/pnas.2023952118

59. Mitchell LA, McCulloch LH, Pinglay S, Berger H, Bosco N, Brosh R, et al. De novo assembly and delivery to mouse cells of a 101 kb functional human gene. Genetics. 2021;218. doi:10.1093/genetics/iyab038

60. Mallory BJ, Tullius TW, Biar CG, Gustafson JA, Bohaczuk SC, Dubocanin D, et al. Principles and functional consequences of plasmid chromatinization in mammalian cells. bioRxiv. 2025. doi:10.1101/2025.05.27.656122

61. Cheng J, Novati G, Pan J, Bycroft C, Žemgulytė A, Applebaum T, et al. Accurate proteome-wide missense variant effect prediction with AlphaMissense. Science. 2023;381: eadg7492.

62. Folta A, Sedeño Cortés AE, Gupta P, McEwen AE, Kao EY, Horike-Pyne M, et al. Imprecision medicine: Systematic gaps in reporting variants of uncertain significance (VUS) and their reclassifications. Genet Med. 2025;27: 101501.

63. Wang C, Ward ME, Chen R, Liu K, Tracy TE, Chen X, et al. Scalable Production of iPSC-Derived Human Neurons to Identify Tau-Lowering Compounds by High-Content Screening. Stem Cell Reports. 2017;9: 1221–1233.

64. Xie Z, Bailey A, Kuleshov MV, Clarke DJB, Evangelista JE, Jenkins SL, et al. Gene Set Knowledge Discovery with Enrichr. Curr Protoc. 2021;1: e90.

65. Schubach M, Maass T, Nazaretyan L, Röner S, Kircher M. CADD v1.7: using protein language models, regulatory CNNs and other nucleotide-level scores to improve genome-wide variant predictions. Nucleic Acids Res. 2024;52: D1143–D1154.

66. Ioannidis NM, Rothstein JH, Pejaver V, Middha S, McDonnell SK, Baheti S, et al. REVEL: An Ensemble Method for Predicting the Pathogenicity of Rare Missense Variants. Am J Hum Genet. 2016;99: 877–885.

67. Frazer J, Notin P, Dias M, Gomez A, Min JK, Brock K, et al. Disease variant prediction with deep generative models of evolutionary data. Nature. 2021;599: 91–95.

68. Pejaver V, Urresti J, Lugo-Martinez J, Pagel KA, Lin GN, Nam H-J, et al. Inferring the molecular and phenotypic impact of amino acid variants with MutPred2. Nat Commun. 2020;11: 5918.

69. Friedman CE, Fayer S, Pendyala S, Chien W-M, Loiben A, Tran L, et al. CRaTER enrichment for on-target gene editing enables generation of variant libraries in hiPSCs. J Mol Cell Cardiol. 2023;179: 60–71.

70. Cardioids reveal self-organizing principles of human cardiogenesis. Cell. 2021;184: 3299–3317.e22.

71. Schmidt C, Deyett A, Ilmer T, Haendeler S, Torres Caballero A, Novatchkova M, et al. Multi-chamber cardioids unravel human heart development and cardiac defects. Cell. 2023;186: 5587–5605.e27.

72. Pejaver V, Byrne AB, Feng B-J, Pagel KA, Mooney SD, Karchin R, et al. Calibration of computational tools for missense variant pathogenicity classification and ClinGen recommendations for PP3/BP4 criteria. Am J Hum Genet. 2022;109: 2163–2177.

73. Wang C, Ward ME, Chen R, Liu K, Tracy TE, Chen X, et al. Scalable Production of iPSC-Derived Human Neurons to Identify Tau-Lowering Compounds by High-Content Screening. Stem Cell Reports. 2017;9: 1221–1233.

